# Rapid antidepressant effects of psilocybin without increased SV2A binding: baseline binding predicts response

**DOI:** 10.64898/2026.11.06.26367247

**Authors:** Hampus Yngwe, Pontus Plavén-Sigray, Max Andersson, Martin Bolin, Maria Beckman, Carl-Johan Ekman, Mikael Tiger, Johan Lundberg

## Abstract

We report a pre-registered [^11^C]UCB-J PET sub-study of a recently published randomized trial in which a single psilocybin dose produced rapid and sustained antidepressant effects in major depressive disorder (MDD). Synaptic loss has been implicated in MDD, and psychedelics induce synaptic remodeling in preclinical models within 24 hours, raising the possibility that psilocybin’s antidepressant effects involve increased synaptic density. We used the PET radioligand [^11^C]UCB-J targeting synaptic vesicle glycoprotein 2A (SV2A) to test this in thirty-five participants with moderate-to-severe recurrent MDD randomized 1:1 to a single dose of 25 mg psilocybin or 100 mg niacin placebo, both with psychotherapeutic support. [^11^C]UCB-J PET was performed 2–7 days before and 15 (±7) days after dosing. Despite significant clinical improvement in the psilocybin group, we found no evidence of a group-level increase in SV2A binding (*V_T_* or *BP_ND_*) relative to niacin in the hippocampus or frontal cortex. In secondary analyses, higher baseline *V_T_* was associated with greater subsequent clinical improvement in the psilocybin group; this survived FDR correction at Day 8 and Day 15 (two of three pre-registered timepoints). Change in *V_T_* was associated with greater subsequent clinical improvement in unadjusted analyses, but these associations were attenuated after adjustment for baseline *V_T_* and baseline MADRS and did not survive FDR correction. These findings indicate that substantial antidepressant improvement after psilocybin can occur without a detectable group-level increase in SV2A binding, while raising the possibility that baseline SV2A binding could help identify individuals more likely to respond.

## Introduction

Synaptic plasticity contributes to resilience and adaptation throughout life (1–5), whereas synaptic dysfunction has been implicated in a range of neurological and psychiatric conditions (6–8), including major depressive disorder (MDD).

The hypothesis that depression involves stress-related synaptopathy in corticolimbic regions is supported by rodent models and postmortem findings of individuals with MDD, which have shown synaptic loss and reduced expression of synapse-related genes (9,10). Such alterations may disrupt neural signaling and network function in circuits implicated in depression(11,12).

The serotonergic psychedelic psilocybin has demonstrated rapid antidepressant effects in randomized clinical trials, that seem to persist well beyond drug elimination (13–15). Whereas acute psychedelic effects are thought to involve serotonin_2A_ (5-HT_2A_) receptor-mediated changes in cortical and network function (16,17), persistent antidepressant effects suggest engagement of downstream processes, including transcriptional regulation and protein synthesis. Psilocin, the active metabolite of psilocybin, has also been reported to bind directly to the brain-derived neurotrophic factor (BDNF) receptor TrkB, suggesting a TrkB/BDNF-dependent pathway for psychedelic-induced plasticity (18). In preclinical models, psychedelics have been shown to promote structural plasticity, including dendritic spine density within 24 hours after administration of psychedelics (19). Psilocybin-induced increases in spine density appear most pronounced during the first days, although a subset of newly formed spines has been reported to persist for at least one month (20). This indicates that a brief pharmacological trigger can result in both rapid and sustained structural synaptic adaptations, mirroring the clinical antidepressant effects observed after a single-dose psilocybin in humans (13,21).

There is ongoing debate regarding how synaptic alterations in depression are to be interpreted. One simplified “loss-restoration” framework proposes that depression is associated with synaptic loss, whereas antidepressants can promote their restoration. In contrast, perspectives such as iPlasticity suggest that treatment may induce a permissive state of heightened plasticity, in which environmental factors shape synaptic remodelling and clinical outcomes (22,23). Taken together, this debate leaves unresolved whether treatment-related plasticity manifests as an increase in synaptic density, and when such changes emerge in relation to treatment and clinical improvement.

Synaptic density has historically been quantified using postmortem and in vitro approaches. The PET radioligand [^11^C]UCB-J, targeting the synaptic vesicle glycoprotein 2A (SV2A), a presynaptic protein widely distributed throughout the brain, has shown close correspondence with the established synaptic marker synaptophysin, enabling estimation of synaptic density in the living human brain (24–26). A landmark study reported lower SV2A binding in affective regions in more severely depressed individuals compared to healthy controls, and that lower binding was associated with increased severity (27). Psilocybin has been shown to increase SV2A binding in the pig brain (28), but whether similar changes occur in humans with MDD, and with a similar time course, remains unknown.

Treatment response in MDD is highly heterogeneous, and the ability to determine in advance which individual is likely to benefit from a given treatment remains limited (29). Pretreatment biomarkers could inform treatment selection and reduce delays associated with ineffective treatment trials. In our previous work, we found that SV2A binding was associated with illness chronicity in MDD, suggesting that SV2A may be linked to illness course and prognosis(30). Moreover, in a study of the rapid-acting antidepressant ketamine, lower baseline SV2A binding was linked to greater subsequent increases in SV2A in prefrontal regions, which in turn related to clinical improvement in exploratory analyses (31).

This raises the question of whether SV2A-binding can predict treatment response to psilocybin.

Against this background, we examined [^11^C]UCB-J PET data from a recently published randomized clinical trial in which psilocybin produced rapid and sustained antidepressant effects in patients with MDD (32). We preregistered the primary hypothesis that psilocybin would increase SV2A binding in the hippocampus and frontal cortex relative to placebo, reflecting a net increase in synaptic density by the time of follow-up PET. In preregistered secondary analyses, we examined whether treatment-related changes in SV2A binding were associated with antidepressant response, and whether baseline SV2A binding was associated with subsequent clinical improvement.

## Methods

### Study Overview

The trial was conducted at Northern Stockholm Psychiatric Clinic and the PET Centre at Karolinska Institutet from January 2021 to February 2024(32).

The protocol was approved by the Swedish Ethical Review Authority (Dnr 2020-03700) and the Swedish Medical Products Agency (Dnr 5.1-2020-57521). Written informed consent was obtained before any study procedures, which were performed in accordance with the Helsinki Declaration and Good Clinical Practice (GCP). The trial protocol is provided as a separate supplementary file. ClinicalTrials.gov ID is NCT04630964.

## Participants

Thirty-five men and women aged 20-65 years and meeting the ICD-10 criteria of an episode of recurrent moderate to severe MDD (Montgomery Åsberg Depression Rating Scale (MADRS) >22 points) were recruited through advertising on social media platforms **(Table 1)** (32). Applicants with a history of psychosis, substance abuse or acute suicidality were excluded, as were applicants with a range of somatic conditions (complete inclusion/exclusion criteria found on Clinicaltrials.gov). Participants were required to remain free of depression treatment throughout the first six weeks of follow-up.

**Table 1.**
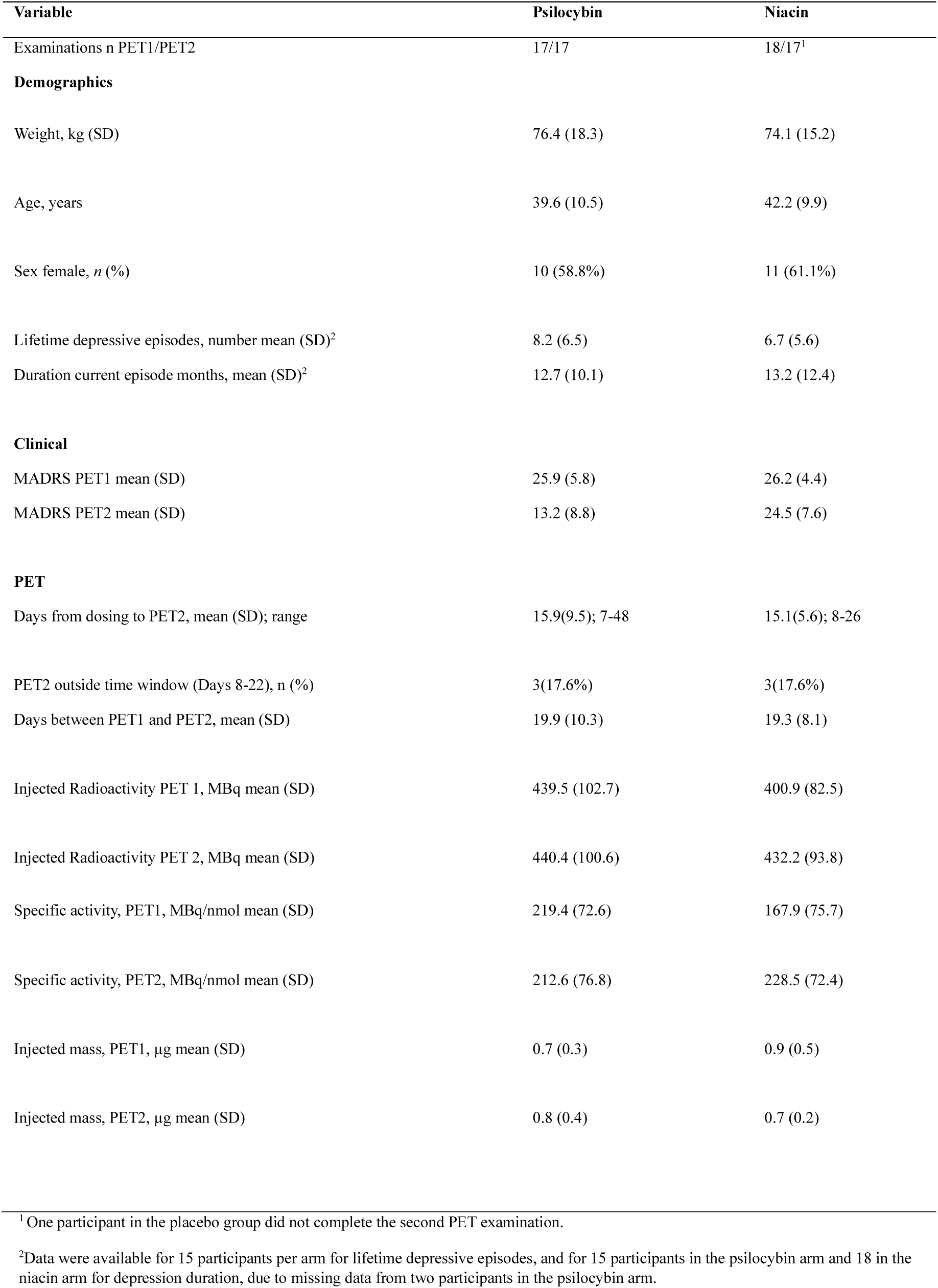
Demographics, Clinical and PET parameters.

| **Variable** | **Psilocybin** | **Niacin** |
|---|---|---|
| Examinations n PET1/PET2 | 17/17 | 18/17<sup>1</sup> |
| **Demographics** |  |  |
| Weight, kg (SD) | 76.4 (18.3) | 74.1 (15.2) |
| Age, years | 39.6 (10.5) | 42.2 (9.9) |
| Sex female, *n* (%) | 10 (58.8%) | 11 (61.1%) |
| Lifetime depressive episodes, number mean (SD)<sup>2</sup> | 8.2 (6.5) | 6.7 (5.6) |
| Duration current episode months, mean (SD)<sup>2</sup> | 12.7 (10.1) | 13.2 (12.4) |
| **Clinical** |  |  |
| MADRS PET1 mean (SD) | 25.9 (5.8) | 26.2 (4.4) |
| MADRS PET2 mean (SD) | 13.2 (8.8) | 24.5 (7.6) |
| **PET** |  |  |
| Days from dosing to PET2, mean (SD); range | 15.9(9.5); 7-48 | 15.1(5.6); 8-26 |
| PET2 outside time window (Days 8-22), n (%) | 3(17.6%) | 3(17.6%) |
| Days between PET1 and PET2, mean (SD) | 19.9 (10.3) | 19.3 (8.1) |
| Injected Radioactivity PET 1, MBq mean (SD) | 439.5 (102.7) | 400.9 (82.5) |
| Injected Radioactivity PET 2, MBq mean (SD) | 440.4 (100.6) | 432.2 (93.8) |
| Specific activity, PET1, MBq/nmol mean (SD) | 219.4 (72.6) | 167.9 (75.7) |
| Specific activity, PET2, MBq/nmol mean (SD) | 212.6 (76.8) | 228.5 (72.4) |
| Injected mass, PET1, µg mean (SD) | 0.7 (0.3) | 0.9 (0.5) |
| Injected mass, PET2, µg mean (SD) | 0.8 (0.4) | 0.7 (0.2) |

<sup>1</sup> One participant in the placebo group did not complete the second PET examination.
<sup>2</sup>Data were available for 15 participants per arm for lifetime depressive episodes, and for 15 participants in the psilocybin arm and 18 in the niacin arm for depression duration, due to missing data from two participants in the psilocybin arm.

## Study Design

Participants were randomized 1:1 to a single dose of 25mg psilocybin or 100mg niacin placebo, both treatment arms received five sessions of psychotherapeutic support from day −1 to day 15. Clinical follow-up extended to 365 days after dosing.

### Magnetic Resonance Imaging

Structural MRI was acquired 7–35 days before dosing at the Solna MRI Centre, Department of Clinical Neuroscience, Karolinska Institutet. The first 26 participants were scanned on a 3-Tesla General Electric Discovery MR750 scanner, and the remaining 9 on a 3-Tesla GE SIGNA Premier scanner. Scanner allocation was balanced across arms.

### Positron Emission Tomography

Participants underwent [^11^C]UCB-J PET (High-Resolution Research Tomograph; Siemens Molecular Imaging) at two timepoints: baseline, 2–7 days before dosing, and follow-up, targeted to 15 days after dosing with an allowable window of ±7 days. PET variables are summarized in **Table 1**. During scanning, participants were positioned supine, with the head fixated.

[^11^C]UCB-J was synthesized at the Karolinska PET centre in accordance with Good Manufacturing Practice (25). A bolus injection containing approximately 400 MBq (±10%, for a participant of 70 kg) was injected via antecubital venous cannula, followed by a saline flush.

A 6-min transmission scan using a rotating ¹³⁷Cs source was acquired for attenuation correction, followed by a 90-min emission scan acquired in list mode. Dynamic images were reconstructed into 37 frames (8 × 15 s, 8 × 30 s, 4 × 60 s, 5 × 120 s, 10 × 300 s, 2 × 600 s) using OP-3D-OSEM with point spread function modelling, yielding a resolution of approximately 2 mm at the center of the field of view.

Arterial blood was sampled via a radial artery cannula for derivation of the arterial input function (AIF) and radiometabolite correction. Continuous sampling was performed during the first 10 minutes using an Automatic Blood Sampling System (ABSS, Alogg technologies, Mariefred, Sweden) followed by discrete manual samples. The parent fraction in plasma and its radiometabolite profile were determined using protein precipitation followed by reversed-phase radio-HPLC.

Head motion was estimated using realignment of all frames to a reference formed from the first frames (first 4 min) in SPM. Four participants had translational displacement > 4 mm in at least one scan, for which motion correction was performed by frame-wise coregistration to the corresponding transmission image and retained if it resulted in changes > 5% in regional outcome estimates and applied to the corresponding paired scan as well.

## Image Processing and Pharmacokinetic Modelling

T1-weighted MR images were segmented into regions of interest (ROIs) using FreeSurfer 6.0 (33) according to the Desikan–Killiany atlas (34). MR images were coregistered to a summed PET image, with ROI definitions transferred to PET using the resulting coregistration matrix. Image preprocessing was done using SPM12 (35), implemented in MATLAB R2014b (The MathWorks Inc., Natick, MA, USA). Time activity curves were extracted for all ROIs.

Kinetic modelling was performed using the open-source R package *kinfitr* (v0.8.0) (36).

Binding was quantified in two ways: (i) total distribution volume (*V_T_*) using the 1-Tissue-Compartment Model with a metabolite corrected arterial input function, and (ii) non-displaceable binding potential (*BP_ND_*) using a simplified reference tissue model (SRTM2) with centrum semiovale as reference region and k2’ estimated from SRTM1 (37–39). All image processing and kinetic modelling were conducted blinded to treatment allocation, using randomized study identifiers.

### Clinical and psychological data

Depressive symptoms were assessed using the MADRS, by blinded raters. Assessments were performed at baseline (PET1), follow-up PET (PET2), and on days 8, 42, and 365 post-dose.

Acute subjective drug effects were assessed using the Mystical Experience Questionnaire (MEQ-30) and the Scale for Psychedelic Intensity Rating (SPIR). SPIR ratings were obtained approximately 90 minutes after dosing, while MEQ-30 was administered at the end of the dosing session.

Challenging Experience Questionnaire (CEQ) and Emotional Breakthrough Inventory (EBI) were used to assess challenging psychological reactions to psychedelics. They were administered at the end of the dosing session. The Lasting Effects Questionnaire (LEQ) is a fusion of questions from CEQ, MEQ and EBI, measuring lasting effects of psychedelic experience, administered at day 8, 15, 42 and 365.

Psychological flexibility was assessed using the Acceptance and Action Questionnaire–II (AAQ-II). AAQ-II was administered during treatment sessions and at follow-up assessments at days 42 and 365. The Satisfaction with Life Scale (SWLS) and Meaning in Life Questionnaire (MLQ) assessed subjective well-being and meaning in life, respectively, and were administered during treatment sessions and at Days 42 and 365.

## Statistical Analysis

### Primary analysis

The statistical analysis was preregistered on aspredicted.org (#152,777) on 28 November 2023, after completion of PET and MRI data collection (spring 2023) but before any statistical analysis of the imaging data; deviations from the registered plan are listed in Supplementary Table 16.

The dependent variable was [^11^C]UCB-J binding, and the primary regions of interest (ROIs) were the bilateral hippocampus (HIP) and frontal cortex (FC). For each outcome measure (i.e. *V_T_* and *BP_ND_*) and ROI we fitted an ANCOVA model with PET2 as the dependent variable, treatment group as predictor and the corresponding PET1 value as covariate. Both *V_T_* and *BP_ND_* were preregistered outcome measures. *V_T_* was emphasized because it is estimated using an arterial input function and does not rely on a reference region, thereby avoiding assumptions regarding stability of the centrum semiovale across treatment conditions; *BP_ND_* was reported as a complementary reference-tissue measure. Analyses were conducted in three steps. In **Model 1** we adjusted for PET1 only; in **Model 2** we additionally included time from dosing to PET2 (days); and in **Model 3** we further adjusted for age and clinical covariates (duration of current episode, baseline depression severity, and number of previous episodes).

The significance level was set to 0.05. Given the high interregional correlations for the binding outcomes, no correction for multiple testing was applied.

Outlier detection, quality control and all analyses were conducted blinded to treatment allocation, using randomized study identifiers.

### Secondary and exploratory analyses

Secondary analyses examined whether change in [^11^C]UCB-J binding from baseline to follow-up examination was associated with change in depressive symptom severity on the MADRS. For the present PET sub study, the MADRS assessment obtained at the same visit as PET2 (Day 15 ±7) was prespecified as the primary clinical endpoint for analyses relating change in PET binding to clinical change. Other follow-up assessments were days 8, 42 and 365. For analyses involving MADRS outcomes at Day 42 and Day 365, observations obtained after post-randomization psychedelic use or reinitiation of antidepressant treatment were censored.

To account for multiple testing in the secondary analyses, false discovery rate (FDR) correction using the Benjamini–Hochberg procedure was applied within each analysis family. For the preregistered correlation analyses, correction was performed separately within each treatment arm and clinical follow-up timepoint across the four regional binding measures (frontal cortex and hippocampus; *V_T_* and *BP_ND_*). For treatment-interaction analyses, FDR correction was applied across the eight interaction tests comprising two regions and four follow-up timepoints. Additional post hoc analyses examining the Δ*V_T_*–ΔMADRS association after adjustment for baseline *V_T_* were treated as a separate analysis family, with FDR correction across the eight tests comprising two regions and four follow-up timepoints within each treatment arm. Nominal and FDR-adjusted *p* values are reported throughout. Pearson’s correlation coefficients (r) were calculated between regional Δ*V_T_* (PET2 – PET1) and ΔMADRS (follow-up – baseline, negative values indicate improvement). Parallel analyses for Δ*BP_ND_* are reported in **Supplementary Tables 2-3**. Analyses were conducted separately within each treatment arm. To assess whether the association between Δ*V_T_* and ΔMADRS differed between treatment groups, we additionally fitted linear models with ΔMADRS as the dependent variable and Δ*Vt*, treatment, and Δ*Vt* × treatment as independent variables. The interaction term was interpreted as the formal test of between-group difference in associations. Because baseline *Vt* was itself associated with clinical outcome and is mathematically incorporated in Δ *Vt*, we performed additional post hoc analyses to assess whether the Δ*V*T–ΔMADRS association was independent of baseline binding. Partial Pearson correlations between Δ*V*T and ΔMADRS were calculated controlling for baseline *Vt*. We additionally fitted linear models including baseline MADRS, baseline *Vt*, and Δ*Vt* as predictors of ΔMADRS. Variance inflation factors were examined to assess collinearity between baseline *Vt* and ΔV*t*. These analyses were considered exploratory robustness analyses. Fully interacted models, including treatment interactions with baseline MADRS, baseline *Vt*, and Δ*Vt*, were additionally fitted to assess whether the adjusted Δ*Vt*–ΔMADRS association differed between treatment groups.

We also examined whether baseline [^11^C]UCB-J binding was associated with antidepressant response on the same timepoints. Pearson’s correlations were calculated between baseline regional binding and ΔMADRS in both treatment arms. In addition, linear models including baseline binding, treatment, and their interaction were fitted to test whether these associations differed between groups. For baseline *V_T_* × treatment interaction analyses, FDR correction was applied across the eight tests comprising two regions and four follow-up timepoints.

Post hoc ROC analyses were performed in the psilocybin group using the response criterion specified in the statistical analysis plan (≥50% reduction in MADRS or >10-point reduction from baseline). AUC estimates and 95% confidence intervals were obtained using stratified bootstrap resampling (10,000 iterations). As a post hoc robustness analysis, associations between baseline *V_T_* and ΔMADRS were additionally adjusted for baseline MADRS to assess whether they were independent of initial depression severity.

Exploratory Spearman rank correlations were conducted to examine associations between regional Δ*V_T_* and Δ*BP_ND_* and psychological outcome measures, including MEQ total score and subscales, SPIR, CEQ, EBI, as well as change in AAQ-II, LEQ, MLQ and SWLS at the available follow-up timepoints.

Finally, we repeated the same ANCOVA model used in the primary analysis across an extended set of anatomically defined ROIs that have previously been implicated in the neurobiology of MDD, including the anterior cingulate and medial prefrontal cortices, insula, amygdala, thalamus, and striatal regions, as well as global grey matter. These analyses were hypothesis-generating in nature.

## Results

All participants in the psilocybin arm completed both PET examinations, while one participant in the niacin arm did not complete the follow-up scan. *V_T_* could not be quantified for two participants in the psilocybin arm, due to incomplete blood sampling.

### Primary analysis

In the PET sample, MADRS decreased from 25.9 to 13.2 in the psilocybin group and from 26.2 to 24.5 in the niacin group between PET1 and PET2 **(Table 1)**. Primary analyses revealed no evidence of a group-level change in SV2A binding two weeks after treatment, as detailed below. We found no evidence for treatment effect on *V_T_* in frontal cortex (psilocybin vs niacin: β = −0.49, 95% CI −2.42 to 1.44, p = 0.61) or hippocampus (β = 0.05, 95% CI −1.39 to 1.50, p = 0.94). Likewise, *BP_ND_* did not differ between groups at follow-up, either in frontal cortex (β = 0.010, 95% CI −0.22 to 0.24, p = 0.93) or hippocampus (β = 0.013, 95% CI −0.14 to 0.16, p = 0.86) **(Figure 1)**. Results were consistent in sensitivity analyses additionally adjusting for time from dosing to PET2 (Model 2) and age and clinical covariates (Model 3) **(Supplementary Table 1)**.

**Figure 1.**
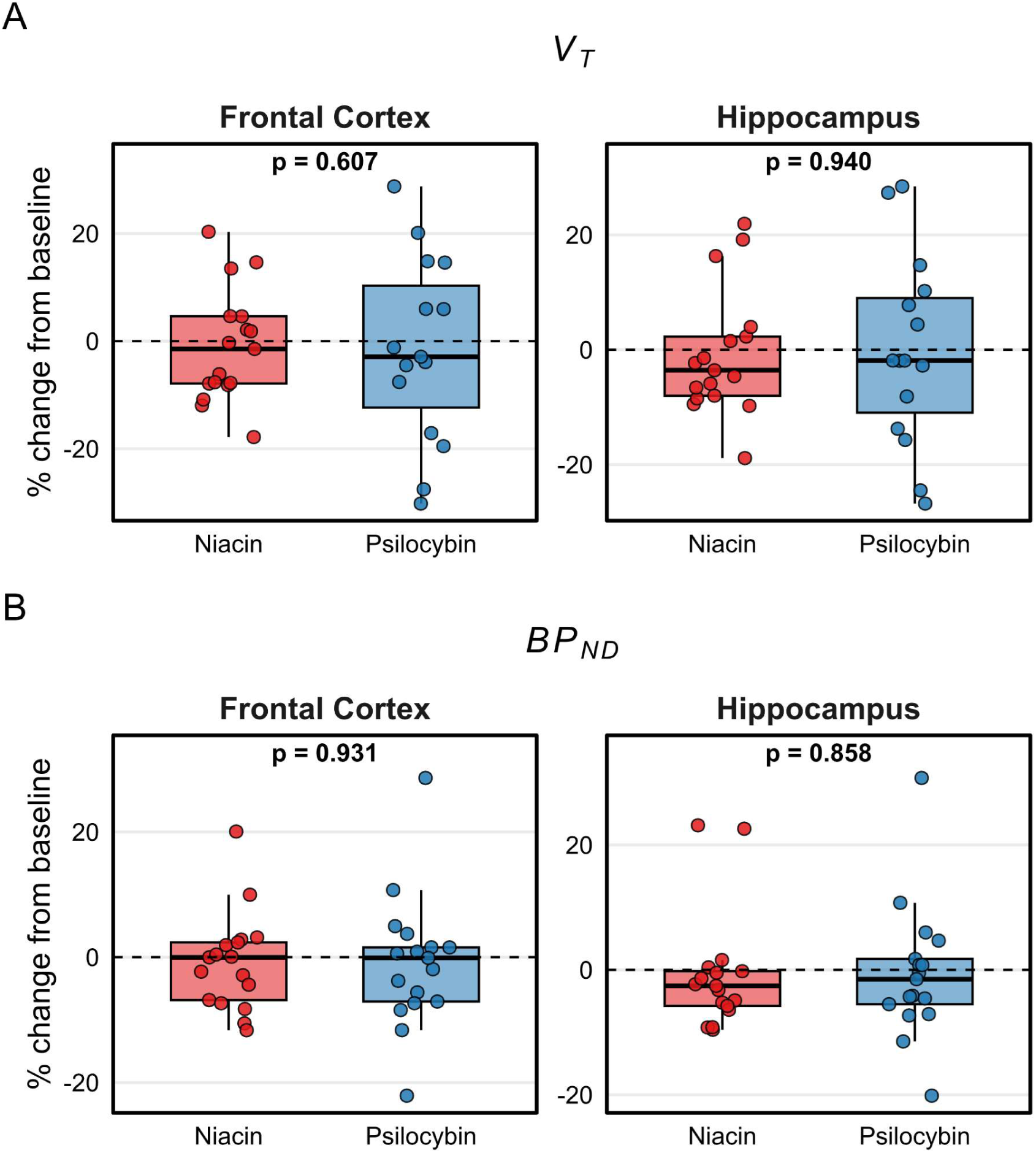
Change from baseline to follow-up in *V_T_* and *BP_ND_* in primary regions of interest by treatment group. Percentage change from baseline to follow-up PET in (A) total distribution volume (*V_T_*) and (B) binding potential relative to non-displaceable uptake (BP_ND_), shown for frontal cortex and hippocampus in the psilocybin and niacin groups. Boxes represent the interquartile range with median; points indicate individual participants. Dashed lines denote no change from baseline. Between-group comparisons were performed using ANCOVA with adjustment for baseline PET (step 1); p-values shown are for the treatment effect (two-sided)

## Secondary Analyses

### Change in [¹¹C]UCB-J binding and antidepressant response

In the unadjusted analyses, lower Δ*V_T_* was associated with greater antidepressant improvement in the psilocybin group, with positive Δ*V_T_*–ΔMADRS correlations across follow-up. The strongest associations were observed at Day 42 (FC: *r* = 0.71, *p* = 0.003, *p*FDR = 0.012; HIP: *r* = 0.67, *p* = 0.007, *p*FDR = 0.013), whereas associations in the niacin group were weak or generally in the opposite direction **(Figure 2, Supplementary Tables 2– 3).** Formal treatment interaction tests did not survive FDR correction **(Supplementary Table 4).** Because baseline *V_T_* was itself associated with clinical improvement and is mathematically contained in Δ*V_T_*, we examined the dependence of Δ*V_T_* on baseline *V_T_* and repeated the analyses adjusting for baseline values. Baseline *V_T_* was negatively correlated with Δ*V_T_* in the psilocybin group (FC: *r* = −0.52; HIP: *r* = −0.39), but this relationship was stronger in the niacin group (FC: *r* = −0.81; HIP: *r* = −0.78), with no evidence that the baseline *V_T_*–Δ*V_T_* relationship differed between treatment groups (FC interaction *p* = 0.823; HIP *p* = 0.480) **(Supplementary Table 7)**. Variance inflation factors were low in the psilocybin group (approximately 1.3–1.5), indicating no substantial multicollinearity between baseline *V_T_* and Δ*V_T_*.

**Figure 2.**
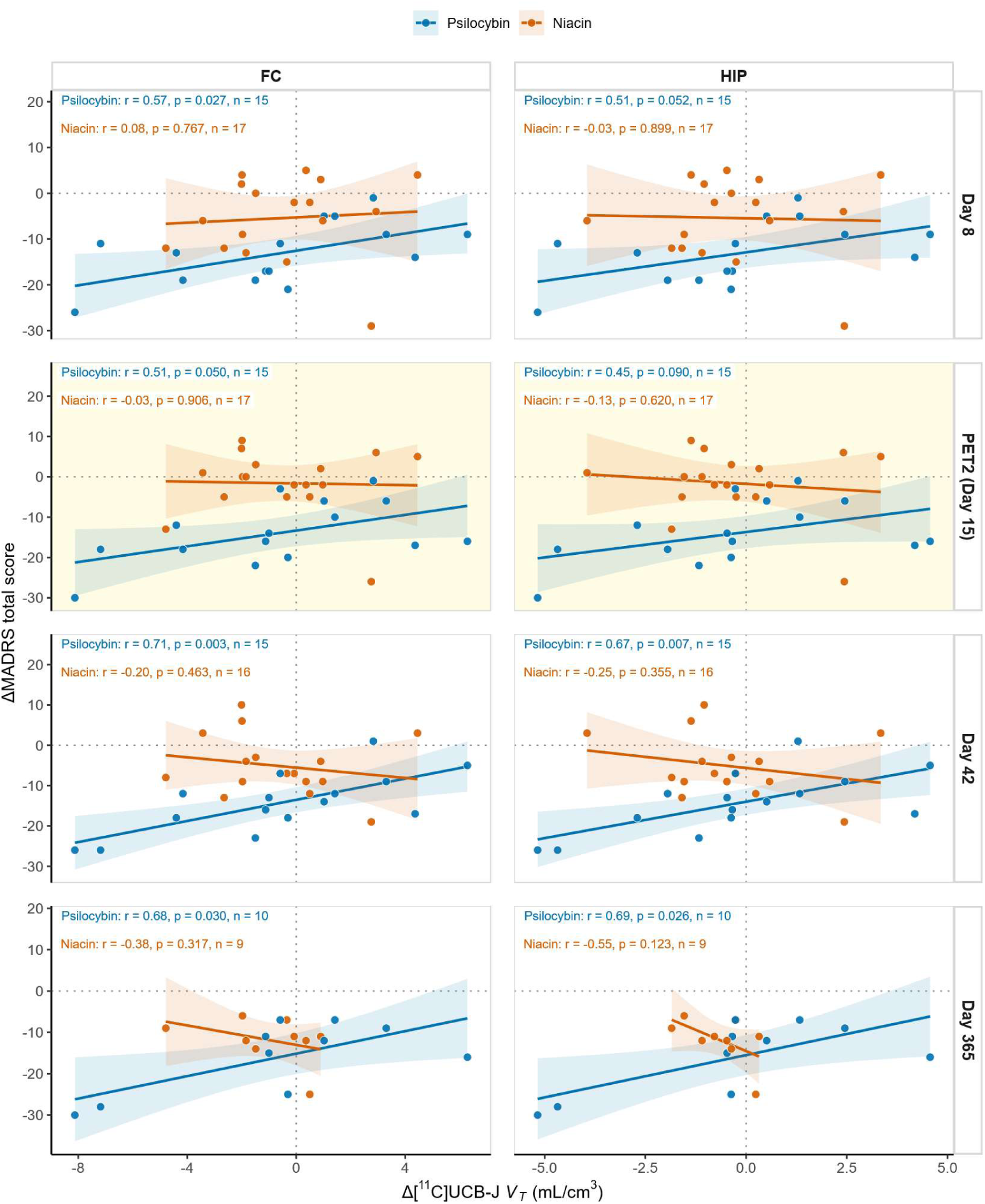
Associations between changes in regional [¹¹C]UCB-J binding and changes in depressive symptom severity across follow-up by treatment group. Associations between change in regional [¹¹C]UCB-J volume of distribution (ΔVₜ; PET2–PET1) and change in depressive symptom severity (ΔMADRS) in the psilocybin (blue) and niacin (red) groups. Observations after antidepressant initiation or psychedelic use outside the study protocol were censored. Panels show frontal cortex (FC) and hippocampus (HIP) at Day 8, PET2 (Day 15), Day 42, and Day 365. Each point represents one participant. Solid lines indicate linear regression fits with 95% confidence intervals, and dotted lines indicate zero change. Pearson correlation coefficients (r), two-sided p values, and sample sizes (n) are provided for each treatment group within each panel; n varies owing to censoring. The predefined PET follow-up timepoint (PET2, Day 15) is shaded for emphasis. Correlations are unadjusted. Abbreviations: ΔMADRS, change in Montgomery–Åsberg Depression Rating Scale score; ΔV_T_, change in volume of distribution; FC, frontal cortex; HIP, hippocampus; PET, positron emission tomography.

Partial correlations adjusting for baseline *V_T_* showed attenuated Δ*V_T_*–ΔMADRS associations **(Supplementary Table 8)**. In regression models additionally adjusting for baseline MADRS, positive associations at Day 42 remained nominally significant in the psilocybin group (FC: β = 1.11, 95% CI 0.09–2.12, p = 0.035; HIP: β = 1.47, 95% CI 0.09–2.86, p = 0.039), but neither survived FDR correction (pFDR = 0.156). No adjusted Δ*V_T_* association survived FDR correction at any time point **(Supplementary Table 9)**. Adjusted interaction analyses provided no robust evidence that the Δ*V_T_*–ΔMADRS association differed between treatment groups **(Supplementary Table 10)**. No corresponding pattern was observed for Δ*BP_ND_*.

### Baseline [¹¹C]UCB-J binding and subsequent antidepressant response

Baseline *V_T_* (PET1) correlated with antidepressant response in the psilocybin arm across follow-up, with higher baseline *V_T_* in both FC and HIP associated with greater symptom improvement at Day 8 (FC: r = −0.63, p = 0.011, p_FDR = 0.026; HIP: r = −0.63, p = 0.013, p_FDR = 0.026) and at the prespecified PET2 endpoint (Day 15; FC: r = −0.61, p = 0.016, p_FDR = 0.032; HIP: r = −0.65, p = 0.009, p_FDR = 0.032). Associations remained in the same direction but did not survive FDR correction at Day 42 (FC: r = −0.51, p = 0.054, p_FDR = 0.108; HIP: r = −0.54, p = 0.038, p_FDR = 0.108) and Day 365 (FC: r = −0.60, p = 0.070, p_FDR = 0.139; HIP: r = −0.63, p = 0.050, p_FDR = 0.139) **(Figure 3**). In contrast, the niacin arm showed no consistent baseline *V_T_*–ΔMADRS associations, with correlations close to zero or positive in direction (Day 8 FC: r = −0.10, p = 0.713; HIP: r = 0.001, p = 0.998; Day 15 FC: r = 0.09, p = 0.727; HIP: r = 0.21, p = 0.419; Day 42 FC: r = 0.37, p = 0.160; HIP: r = 0.44, p = 0.088; Day 365 FC: r = 0.17, p = 0.656; HIP: r = 0.36, p = 0.336) **(Supplementary Table 5).** Formal interaction analyses showed nominal treatment-group differences in the association between baseline *V_T_* and antidepressant response at PET2 in hippocampus (interaction β = −2.80, p = 0.024), at Day 42 in frontal cortex (β = −2.28, p = 0.019) and hippocampus (β = −3.15, p = 0.007), and at Day 365 in hippocampus (β = −3.12, p = 0.047). However, none of these interactions survived FDR correction (all pFDR ≥ 0.060) (Supplementary Table 6). No corresponding pattern was observed for baseline *BP_ND_*. In a post hoc robustness analysis adjusting for baseline MADRS, the associations remained nominally significant at Day 8 and PET2 in both frontal cortex and hippocampus (all p < 0.05), although they did not survive FDR correction across the eight robustness tests (pFDR = 0.082) **(Supplementary Table 5)**. Sensitivity analyses excluding the participant with the highest baseline *V_T_* are presented in **Supplementary Table 17**. At PET2, baseline *V_T_* showed an AUC of 0.89 (bootstrap 95% CI 0.66–1.00) in frontal cortex and 0.91 (0.68–1.00) in hippocampus for distinguishing responders from non-responders **(Supplementary Table 18 and Supplementary Figure 1).**

**Figure 3.**
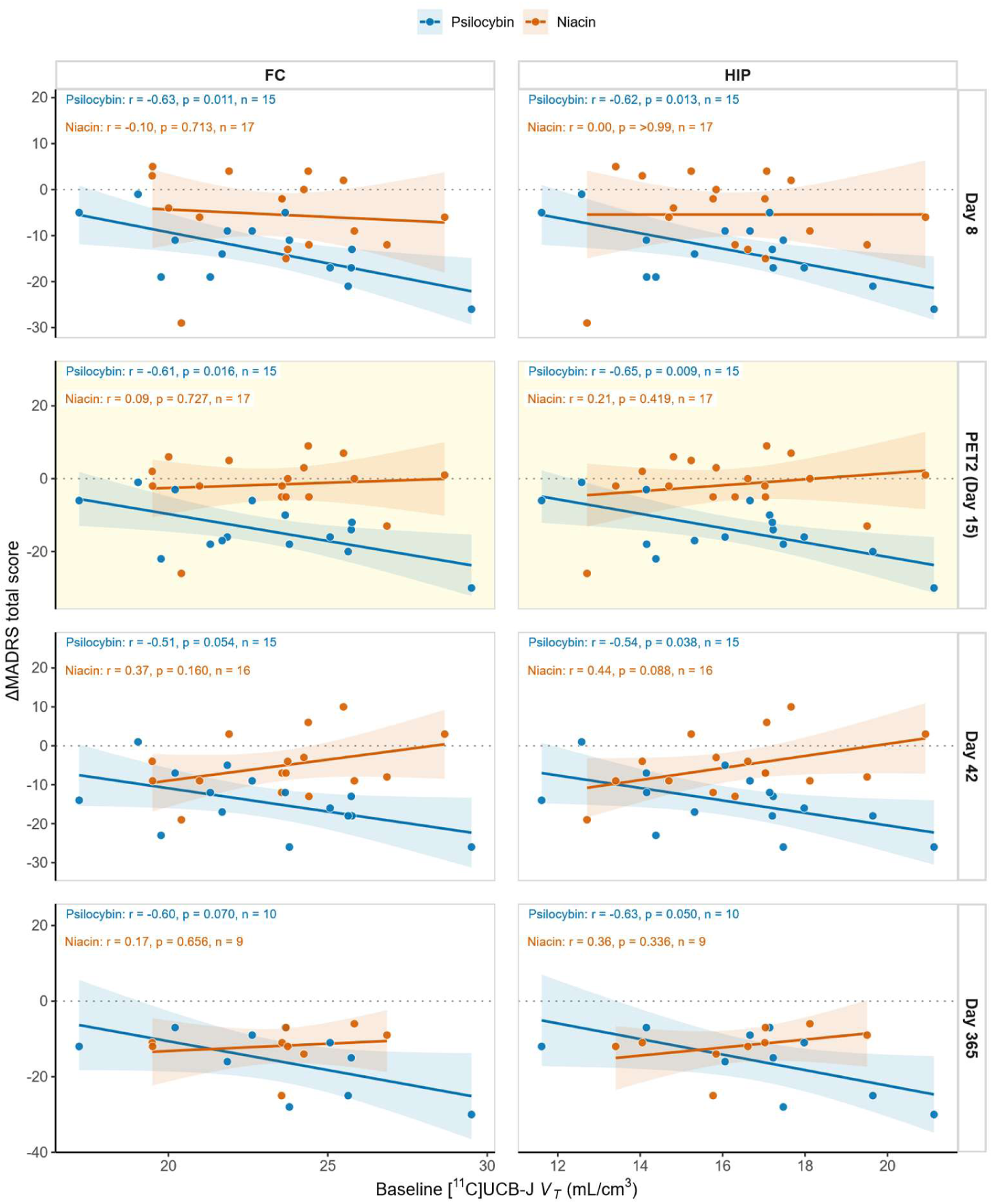
Associations between baseline regional [¹¹C]UCB-J binding and changes in depressive symptom severity across follow-up by treatment group. Associations between baseline regional [¹¹C]UCB-J volume of distribution (V*_T_*; PET1) and change in depressive symptom severity (ΔMADRS) in the psilocybin (blue) and niacin (red) groups. Observations after antidepressant initiation or psychedelic use outside the study protocol were censored. Panels show frontal cortex (FC) and hippocampus (HIP) at Day 8, PET2 (Day 15), Day 42, and Day 365. Each point represents one participant. Solid lines indicate linear regression fits with 95% confidence intervals, and the horizontal dotted line indicates zero change in MADRS. Pearson correlation coefficients (r), two-sided unadjusted p values, and sample sizes (n) are provided for each treatment group within each panel; n varies owing to censoring. False discovery rate (FDR) correction was applied separately within each treatment group. The predefined PET follow-up timepoint (PET2, Day 15) is shaded for emphasis. Abbreviations: ΔMADRS, change in Montgomery–Åsberg Depression Rating Scale score; FC, frontal cortex; HIP, hippocampus; PET, positron emission tomography; V_T_, volume of distribution.

## Exploratory analyses

Exploratory analyses of psychological questionnaire measures yielded largely insignificant results. However, greater reductions in *V_T_* were associated with greater change in LEQ scores at Day 365 in the psilocybin group in both frontal cortex and hippocampus after FDR correction **(Supplementary Table 11)**. Given the large number of exploratory comparisons, these isolated findings should be interpreted cautiously. In the exploratory ROI analyses, no significant treatment effects were observed in any region for either *V_T_* or *BP_ND_*. Effect estimates were small and confidence intervals included zero **(Supplementary Tables 12 and 13**). Descriptive PET binding values for the exploratory regions are provided in **Supplementary Table 15**.

## Discussion

In this placebo-controlled experiment in patients with MDD, we found no evidence that a single 25 mg dose of psilocybin administered with psychotherapeutic support increased synaptic density, indexed by [^11^C]UCB-J binding to SV2A, two weeks after administration, despite rapid and sustained antidepressant effects in the psilocybin group. However, confidence intervals were wide and small-to-moderate treatment effects cannot be excluded. This finding contrasts with a report of increased SV2A binding following psilocybin administration in the pig brain (28), but is consistent with a recent study in healthy volunteers that found no significant overall increase in SV2A binding one week after psilocybin, underscoring that translation to humans remains uncertain(40). Our finding extends this observation to patients with MDD, providing no support for a simple synaptic loss– restoration model of psilocybin’s antidepressant effects. While SV2A PET may not capture all treatment-related synaptic plasticity, the findings are also consistent with the possibility that the antidepressant effects are mediated through other pathways, including expectancy and unblinding effects.

In the secondary analyses, higher baseline SV2A binding was associated with greater subsequent clinical improvement in the psilocybin group, with associations surviving correction for multiple comparisons at Day 8 and PET2 in both regions examined.

Associations remained in the same direction at Day 42 and Day 365 but did not survive FDR correction. One possible interpretation is that higher SV2A binding reflects a greater synaptic reserve that may facilitate treatment response, even if the antidepressant effect itself is not accompanied by an increase in SV2A binding. This finding raises the possibility that SV2A binding is associated with treatment response, but because the treatment interactions did not survive FDR correction it cannot be determined whether this reflects a psilocybin-specific predictor or a general prognostic marker. The ability to identify patients likely to respond could improve the risk-benefit balance of psilocybin treatment as it moves toward clinical use. In a post hoc analysis, we therefore explored its discriminative performance using receiver operating characteristic analyses. At PET2, baseline *V*_T_ showed good discrimination between responders and non-responders, with area under the curve values of 0.89 in frontal cortex and 0.91 in hippocampus. These estimates should be interpreted cautiously given the small sample and the small number of non-responders and require prospective validation.

In contrast, the apparent association between greater reductions in *V_T_* and greater symptom improvement raised the possibility that psilocybin responders might show an early post-treatment reduction in SV2A binding. This was contrary to our hypothesis, but given that SV2A expression seems to generally increase during brain maturation, this could hypothetically reflect transient synaptic remodeling during a period of heightened, juvenile-like plasticity (22,41,42). However, this association was substantially attenuated after adjustment for baseline *V_T_* and baseline MADRS and did not survive correction for multiple comparisons. Fully adjusted interaction models provided no robust evidence that the association differed between treatment groups.

The study has several limitations, including the modest sample size. Missing arterial data prevented estimation of *V_T_* for two participants in the psilocybin group. The *V_T_*–MADRS analyses at Day 365 were based on a reduced sample due to censoring. Plasma free fraction (*f*_P_) was not measured, limiting interpretation of *V_T_*. As a presynaptic marker, SV2A binding may not capture postsynaptic or dendritic plasticity, such as spine remodeling, that could be relevant to treatment response. In addition, the single follow-up PET scan at approximately 15 days provides only a limited temporal snapshot of post-treatment SV2A binding and could miss out on earlier transient or later changes arising downstream of the clinical response.

In conclusion, our findings suggest that psilocybin can exert antidepressant effects in MDD without detectable increases in SV2A binding compared to control, two weeks after treatment. This does not rule out synaptic plasticity following psilocybin treatment but highlights the limitations of SV2A binding as a proxy marker. Secondary findings raise the possibility that SV2A binding may have a predictive value for treatment response. Put together, this suggests a more complex pattern than can be explained by a synaptic loss-restoration narrative. Further clarification of the role of synaptic plasticity in psilocybin’s antidepressant effects would benefit from larger samples, earlier and repeated PET assessments, and multimodal approaches.

## Acknowledgements

We thank the staff at the Karolinska Experimental Research and Imaging Centre (KERIC) and the Brain Stimulation Unit at Northern Stockholm Psychiatry for their contributions to the PET data acquisition and clinical conduct of the study.

We thank Aleksander Lebedev, Kasim Acar and Predrag Petrovic at Karolinska Institutet for their contributions to collecting and sharing MRI-data used in this project.

## Author Contributions

**Conceptualization:** P.P.-S., M.Bec., M.T., and J.L.

**Data acquisition:** H.Y. and C.-J.E.

**PET data processing:** P.P.-S., M.A., and M.Bol.

**Formal analysis:** H.Y., P.P.-S., and J.L.

**Writing – original draft:** H.Y., P.P.-S., and J.L.

**Writing – review and editing:** All authors.

**Supervision:** P.P.-S. and J.L.

All authors reviewed and approved the final manuscript.

## Data Availability

The individual-level data underlying this study are not publicly available due to participant confidentiality and ethical restrictions. Requests for data sharing may be considered on a case-by-case basis, subject to applicable ethical approvals, legal requirements, and institutional data-sharing agreements. Requests should be directed to the corresponding author.

