## Supplementary Appendix for "Rapid antidepressant effects of psilocybin without increased SV2A binding: baseline binding predicts response"

**Contents**


1. **Supplementary Table 1. Treatment effects on SV2A binding across models.**
2. **Supplementary Table 2. Correlations between changes in SV2A binding and depressive symptom severity at PET2.**
3. **Supplementary Table 3. Correlations between changes in SV2A binding and depressive symptom severity at other follow-up timepoints.**
4. **Supplementary Table 4. Between-group differences in the association between Δ$V_t$ and ΔMADRS across follow-up.**
5. **Supplementary Table 5. Associations between baseline SV2A binding and subsequent changes in depressive symptom severity.**
6. **Supplementary Table 6. Treatment interactions in the association between baseline $V_t$ and changes in depressive symptom severity.**
7. **Supplementary Table 7. Relationship between baseline $V_t$ and subsequent change in $V_t$.**
8. **Supplementary Table 8. Partial correlations between Δ$V_t$ and ΔMADRS adjusted for baseline $V_t$.**
9. **Supplementary Table 9. Adjusted within-arm associations between Δ$V_t$ and ΔMADRS.**
10. **Supplementary Table 10. Fully interacted treatment-robustness models for the association between Δ$V_t$ and ΔMADRS.**
11. **Supplementary Table 11. Exploratory associations between PET measures and psychological questionnaire outcomes.**
12. **Supplementary Table 12. Exploratory ANCOVA analyses in additional $V_t$ regions of interest.**
13. **Supplementary Table 13. Exploratory ANCOVA analyses in additional $BP_{ND}$ regions of interest.**
14. **Supplementary Table 14. Descriptive SV2A binding in prespecified primary regions of interest.**
15. **Supplementary Table 15. Descriptive SV2A PET binding in exploratory regions of interest.**
16. **Supplementary Table 16. Deviations from, and additions to, the preregistered analysis plan.**
17. **Supplementary Table 17. Sensitivity of the baseline $V_t$–ΔMADRS association to the participant with the highest baseline $V_t$.**
18. **Supplementary Table 18. ROC analysis of baseline $V_t$ under two clinical response definitions.**
19. **Supplementary ROC methods and results**
20. **Supplementary Figure 1. Baseline [$^{11}$C]UCB-J $V_t$ as a classifier of clinical response at PET2 (Day 15).**

### Supplementary Table 1. Treatment effects on SV2A binding across models.

Estimated treatment effects (psilocybin vs niacin) on regional VT and BPND at PET2 in frontal cortex and hippocampus. Model 1 adjusts for baseline binding; Model 2 additionally adjusts for time from dosing to PET2; Model 3 further adjusts for age and clinical covariates.

| Model | Outcome | ROI | β | SE | 95% CI | p | N | $R^2$ | Adj. $R^2$ |
|---|---|---|---|---|---|---|---|---|---|
| 1 | BPND | Frontal cortex | 0.010 | 0.11 | -0.22 to 0.24 | 0.931 | 34 | 0.36 | 0.32 |
| 2 | BPND | Frontal cortex | 0.013 | 0.12 | -0.22 to 0.25 | 0.914 | 34 | 0.37 | 0.30 |
| 3 | BPND | Frontal cortex | 0.10 | 0.11 | -0.13 to 0.33 | 0.364 | 29 | 0.65 | 0.54 |
| 1 | BPND | Hippocampus | 0.013 | 0.073 | -0.14 to 0.16 | 0.858 | 34 | 0.38 | 0.34 |
| 2 | BPND | Hippocampus | 0.013 | 0.075 | -0.14 to 0.17 | 0.863 | 34 | 0.38 | 0.32 |
| 3 | BPND | Hippocampus | 0.081 | 0.073 | -0.072 to 0.23 | 0.282 | 29 | 0.64 | 0.52 |
| 1 | VT | Frontal cortex | -0.49 | 0.94 | -2.42 to 1.44 | 0.607 | 32 | 0.12 | 0.05 |
| 2 | VT | Frontal cortex | -0.38 | 0.94 | -2.30 to 1.53 | 0.686 | 32 | 0.17 | 0.08 |
| 3 | VT | Frontal cortex | 0.29 | 0.96 | -1.71 to 2.29 | 0.767 | 28 | 0.38 | 0.17 |
| 1 | VT | Hippocampus | 0.053 | 0.71 | -1.39 to 1.50 | 0.940 | 32 | 0.24 | 0.19 |
| 2 | VT | Hippocampus | 0.12 | 0.71 | -1.33 to 1.57 | 0.867 | 32 | 0.26 | 0.18 |
| 3 | VT | Hippocampus | 0.54 | 0.75 | -1.01 to 2.10 | 0.474 | 28 | 0.45 | 0.26 |

Abbreviations: BPND, non-displaceable binding potential; FC, frontal cortex; HIP, hippocampus; PET2, second PET examination; VT, total volume of distribution; β, regression coefficient; CI, confidence interval.

### Supplementary Table 2. Pearson correlations between change in SV2A binding from baseline to PET2 and change in depressive symptom severity.

| Treatment | Measure | ROI | n | r | p | pFDR |
|---|---|---|---|---|---|---|
| Psilocybin | ΔBPND | Frontal cortex | 17 | 0.065 | 0.804 | 0.887 |
| Psilocybin | ΔBPND | Hippocampus | 17 | -0.037 | 0.887 | 0.887 |
| Psilocybin | ΔVT | Frontal cortex | 15 | 0.514 | 0.050 | 0.180 |
| Psilocybin | ΔVT | Hippocampus | 15 | 0.453 | 0.090 | 0.180 |
| Niacin | ΔBPND | Frontal cortex | 17 | -0.203 | 0.434 | 0.826 |
| Niacin | ΔBPND | Hippocampus | 17 | -0.403 | 0.108 | 0.433 |
| Niacin | ΔVT | Frontal cortex | 17 | -0.031 | 0.906 | 0.906 |
| Niacin | ΔVT | Hippocampus | 17 | -0.130 | 0.620 | 0.826 |

Negative ΔMADRS values indicate symptom improvement. pFDR denotes Benjamini–Hochberg FDR correction within treatment arm at PET2 across the four imaging measures (VT/BPND × frontal cortex/hippocampus).

**Supplementary Table 3. Within-group Pearson correlations between change in SV2A binding and change in depressive symptom severity at follow-up timepoints other than PET2.**

| Timepoint | Treatment | Measure | ROI | n | r | p | pFDR |
|---|---|---|---|---|---|---|---|
| Day 8 | Psilocybin | ΔVT | Frontal cortex | 15 | 0.567 | 0.027 | 0.104 |
| Day 8 | Psilocybin | ΔVT | Hippocampus | 15 | 0.511 | 0.052 | 0.104 |
| Day 8 | Psilocybin | ΔBPND | Frontal cortex | 17 | 0.269 | 0.296 | 0.364 |
| Day 8 | Psilocybin | ΔBPND | Hippocampus | 17 | 0.235 | 0.364 | 0.364 |
| Day 8 | Niacin | ΔVT | Frontal cortex | 17 | 0.078 | 0.767 | 0.899 |
| Day 8 | Niacin | ΔVT | Hippocampus | 17 | -0.033 | 0.899 | 0.899 |
| Day 8 | Niacin | ΔBPND | Frontal cortex | 17 | -0.171 | 0.512 | 0.899 |
| Day 8 | Niacin | ΔBPND | Hippocampus | 17 | -0.344 | 0.177 | 0.707 |
| Day 42 | Psilocybin | ΔVT | Frontal cortex | 15 | 0.709 | 0.003 | 0.012 |
| Day 42 | Psilocybin | ΔVT | Hippocampus | 15 | 0.667 | 0.007 | 0.013 |
| Day 42 | Psilocybin | ΔBPND | Frontal cortex | 17 | 0.329 | 0.197 | 0.263 |
| Day 42 | Psilocybin | ΔBPND | Hippocampus | 17 | 0.228 | 0.380 | 0.380 |
| Day 42 | Niacin | ΔVT | Frontal cortex | 16 | -0.198 | 0.463 | 0.822 |
| Day 42 | Niacin | ΔVT | Hippocampus | 16 | -0.248 | 0.355 | 0.822 |
| Day 42 | Niacin | ΔBPND | Frontal cortex | 16 | -0.061 | 0.822 | 0.822 |
| Day 42 | Niacin | ΔBPND | Hippocampus | 16 | -0.106 | 0.697 | 0.822 |
| Day 365 | Psilocybin | ΔVT | Frontal cortex | 10 | 0.682 | 0.030 | 0.060 |
| Day 365 | Psilocybin | ΔVT | Hippocampus | 10 | 0.694 | 0.026 | 0.060 |
| Day 365 | Psilocybin | ΔBPND | Frontal cortex | 11 | 0.137 | 0.688 | 0.865 |
| Day 365 | Psilocybin | ΔBPND | Hippocampus | 11 | 0.058 | 0.865 | 0.865 |
| Day 365 | Niacin | ΔVT | Frontal cortex | 9 | -0.377 | 0.317 | 0.634 |
| Day 365 | Niacin | ΔVT | Hippocampus | 9 | -0.552 | 0.123 | 0.494 |
| Day 365 | Niacin | ΔBPND | Frontal cortex | 9 | 0.015 | 0.970 | 0.970 |
| Day 365 | Niacin | ΔBPND | Hippocampus | 9 | 0.175 | 0.652 | 0.869 |

Day 42 and Day 365 use censored outcomes. Negative ΔMADRS values indicate symptom improvement. pFDR is corrected within treatment arm and follow-up timepoint across the four imaging measures.

### Supplementary Table 4. Between-group differences in the association between ΔVT and ΔMADRS across follow-up.

| Timepoint | ROI | n total | n psilocybin | n niacin | βinteraction | 95% CI | p | pFDR |
|---|---|---|---|---|---|---|---|---|
| Day 8 | Frontal cortex | 32 | 15 | 17 | 0.659 | -1.294 to 2.612 | 0.495 | 0.495 |
| Day 8 | Hippocampus | 32 | 15 | 17 | 1.411 | -1.335 to 4.157 | 0.301 | 0.345 |
| PET2 (Day 15) | Frontal cortex | 32 | 15 | 17 | 1.081 | -0.877 to 3.039 | 0.268 | 0.345 |
| PET2 (Day 15) | Hippocampus | 32 | 15 | 17 | 1.859 | -0.883 to 4.600 | 0.176 | 0.281 |
| Day 42 | Frontal cortex | 31 | 15 | 16 | 1.951 | 0.179 to 3.723 | 0.032 | 0.129 |
| Day 42 | Hippocampus | 31 | 15 | 16 | 2.907 | 0.404 to 5.410 | 0.024 | 0.129 |
| Day 365 | Frontal cortex | 19 | 10 | 9 | 2.536 | -0.271 to 5.342 | 0.073 | 0.147 |
| Day 365 | Hippocampus | 19 | 10 | 9 | 6.139 | 0.021 to 12.257 | 0.049 | 0.132 |

Linear models included ΔVT, treatment, and ΔVT × treatment. Niacin is the reference group; βinteraction is the difference in slope for psilocybin relative to niacin. pFDR is Benjamini–Hochberg corrected across the eight VT interaction tests (2 ROIs × 4 timepoints). Day 42 and Day 365 use censored outcomes.

### Supplementary Table 5. Association between baseline SV2A binding and subsequent change in depressive symptom severity.

#### Panel A. Unadjusted correlations between baseline SV2A binding and subsequent change in depressive symptom severity.

| Timepoint | Treatment | Measure | ROI | n | r | p | pFDR |
|---|---|---|---|---|---|---|---|
| Day 8 | Psilocybin | Baseline VT | Frontal cortex | 15 | -0.633 | 0.011 | 0.026 |
| Day 8 | Psilocybin | Baseline VT | Hippocampus | 15 | -0.625 | 0.013 | 0.026 |
| Day 8 | Psilocybin | Baseline BPND | Frontal cortex | 17 | -0.234 | 0.367 | 0.424 |
| Day 8 | Psilocybin | Baseline BPND | Hippocampus | 17 | -0.208 | 0.424 | 0.424 |
| Day 8 | Niacin | Baseline VT | Frontal cortex | 17 | -0.096 | 0.713 | 0.953 |
| Day 8 | Niacin | Baseline VT | Hippocampus | 17 | 0.001 | >0.99 | >0.99 |
| Day 8 | Niacin | Baseline BPND | Frontal cortex | 17 | -0.165 | 0.527 | 0.953 |
| Day 8 | Niacin | Baseline BPND | Hippocampus | 17 | 0.096 | 0.715 | 0.953 |
| PET2 (Day 15) | Psilocybin | Baseline VT | Frontal cortex | 15 | -0.609 | 0.016 | 0.032 |
| PET2 (Day 15) | Psilocybin | Baseline VT | Hippocampus | 15 | -0.648 | 0.009 | 0.032 |
| PET2 (Day 15) | Psilocybin | Baseline BPND | Frontal cortex | 17 | 0.016 | 0.951 | 0.951 |
| PET2 (Day 15) | Psilocybin | Baseline BPND | Hippocampus | 17 | -0.042 | 0.874 | 0.951 |
| PET2 (Day 15) | Niacin | Baseline VT | Frontal cortex | 17 | 0.091 | 0.727 | 0.943 |
| PET2 (Day 15) | Niacin | Baseline VT | Hippocampus | 17 | 0.210 | 0.419 | 0.838 |
| PET2 (Day 15) | Niacin | Baseline BPND | Frontal cortex | 17 | 0.019 | 0.943 | 0.943 |
| PET2 (Day 15) | Niacin | Baseline BPND | Hippocampus | 17 | 0.442 | 0.075 | 0.302 |
| Day 42 | Psilocybin | Baseline VT | Frontal cortex | 15 | -0.506 | 0.054 | 0.108 |
| Day 42 | Psilocybin | Baseline VT | Hippocampus | 15 | -0.540 | 0.038 | 0.108 |
| Day 42 | Psilocybin | Baseline BPND | Frontal cortex | 17 | 0.095 | 0.717 | 0.956 |
| Day 42 | Psilocybin | Baseline BPND | Hippocampus | 17 | 0.012 | 0.963 | 0.963 |
| Day 42 | Niacin | Baseline VT | Frontal cortex | 16 | 0.369 | 0.160 | 0.319 |
| Day 42 | Niacin | Baseline VT | Hippocampus | 16 | 0.440 | 0.088 | 0.319 |
| Day 42 | Niacin | Baseline BPND | Frontal cortex | 16 | -0.091 | 0.737 | 0.737 |
| Day 42 | Niacin | Baseline BPND | Hippocampus | 16 | 0.241 | 0.368 | 0.491 |
| Day 365 | Psilocybin | Baseline VT | Frontal cortex | 10 | -0.595 | 0.070 | 0.139 |
| Day 365 | Psilocybin | Baseline VT | Hippocampus | 10 | -0.631 | 0.050 | 0.139 |
| Day 365 | Psilocybin | Baseline BPND | Frontal cortex | 11 | 0.376 | 0.255 | 0.340 |
| Day 365 | Psilocybin | Baseline BPND | Hippocampus | 11 | 0.318 | 0.341 | 0.341 |
| Day 365 | Niacin | Baseline VT | Frontal cortex | 9 | 0.173 | 0.656 | 0.787 |
| Day 365 | Niacin | Baseline VT | Hippocampus | 9 | 0.364 | 0.336 | 0.787 |
| Day 365 | Niacin | Baseline BPND | Frontal cortex | 9 | -0.205 | 0.598 | 0.787 |
| Day 365 | Niacin | Baseline BPND | Hippocampus | 9 | 0.106 | 0.787 | 0.787 |

Negative ΔMADRS values indicate symptom improvement; therefore, negative correlations indicate that higher baseline binding is associated with greater symptom improvement. pFDR is corrected within treatment arm and follow-up timepoint across the four baseline imaging measures. Day 42 and Day 365 use censored outcomes. The Day 365 baseline-binding analysis is additional/exploratory relative to the preregistered Day 8, PET2 and Day 42 analyses.

**Panel B. Associations between baseline VT and subsequent change in depressive symptom severity adjusted for baseline MADRS.**

| Timepoint | ROI | n | βbaseline VT | 95% CI | p | pFDR |
|---|---|---|---|---|---|---|
| Day 8 | Frontal cortex | 15 | -1.185 | -2.230 to -0.142 | 0.029 | 0.082 |
| Day 8 | Hippocampus | 15 | -1.454 | -2.830 to -0.081 | 0.040 | 0.082 |
| PET2 (Day 15) | Frontal cortex | 15 | -1.154 | -2.250 to -0.055 | 0.041 | 0.082 |
| PET2 (Day 15) | Hippocampus | 15 | -1.533 | -2.930 to -0.139 | 0.034 | 0.082 |
| Day 42 | Frontal cortex | 15 | -1.066 | -2.390 to 0.261 | 0.106 | 0.141 |
| Day 42 | Hippocampus | 15 | -1.454 | -3.130 to 0.227 | 0.084 | 0.134 |
| Day 365 | Frontal cortex | 10 | -0.894 | -2.460 to 0.676 | 0.220 | 0.220 |
| Day 365 | Hippocampus | 10 | -1.234 | -3.220 to 0.756 | 0.186 | 0.213 |

Models were fitted within the psilocybin arm as ΔMADRS ~ baseline MADRS + baseline VT. βbaseline VT is the coefficient for regional baseline VT after adjustment for baseline MADRS; negative coefficients indicate that higher baseline VT is associated with greater symptom improvement. pFDR is Benjamini–Hochberg corrected across the eight tests (2 ROIs × 4 timepoints). Day 42 and Day 365 use censored outcomes. This analysis was post hoc and was performed as a robustness check because baseline MADRS is mathematically incorporated in ΔMADRS.

### Supplementary Table 6. Treatment interaction in the association between baseline VT and change in depressive symptom severity.

| Timepoint | ROI | n | βinteraction | 95% CI | p | pFDR |
|---|---|---|---|---|---|---|
| Day 8 | Frontal cortex | 32 | -1.025 | -3.009 to 0.960 | 0.299 | 0.299 |
| Day 8 | Hippocampus | 32 | -1.675 | -4.177 to 0.828 | 0.181 | 0.207 |
| PET2 (Day 15) | Frontal cortex | 32 | -1.765 | -3.729 to 0.198 | 0.076 | 0.122 |
| PET2 (Day 15) | Hippocampus | 32 | -2.798 | -5.198 to -0.398 | 0.024 | 0.064 |
| Day 42 | Frontal cortex | 31 | -2.276 | -4.155 to -0.397 | 0.019 | 0.064 |
| Day 42 | Hippocampus | 31 | -3.153 | -5.388 to -0.917 | 0.007 | 0.060 |
| Day 365 | Frontal cortex | 19 | -1.914 | -4.390 to 0.561 | 0.120 | 0.160 |
| Day 365 | Hippocampus | 19 | -3.122 | -6.203 to -0.040 | 0.047 | 0.095 |

Linear models included baseline VT, treatment, and baseline VT × treatment. Niacin is the reference group. pFDR is Benjamini–Hochberg corrected across the eight baseline-VT interaction tests (2 ROIs × 4 timepoints). Day 42 and Day 365 use censored outcomes.

### Supplementary Table 7. Relationship between baseline VT and subsequent change in VT from PET1 to PET2.

#### Panel A. Within-group correlations.

| Treatment | ROI | n | r | 95% CI | p | pFDR |
|---|---|---|---|---|---|---|
| Psilocybin | Frontal cortex | 15 | -0.518 | -0.814 to -0.008 | 0.048 | 0.096 |
| Psilocybin | Hippocampus | 15 | -0.391 | -0.752 to 0.152 | 0.150 | 0.150 |
| Niacin | Frontal cortex | 17 | -0.808 | -0.928 to -0.534 | <0.001 | <0.001 |
| Niacin | Hippocampus | 17 | -0.775 | -0.915 to -0.469 | <0.001 | <0.001 |

#### Panel B. Treatment interaction in the baseline VT–ΔVT relationship.

| ROI | n total | n psilocybin | n niacin | βinteraction | 95% CI | p | pFDR |
|---|---|---|---|---|---|---|---|
| Frontal cortex | 32 | 15 | 17 | 0.077 | -0.621 to 0.775 | 0.823 | 0.823 |
| Hippocampus | 32 | 15 | 17 | 0.227 | -0.424 to 0.879 | 0.480 | 0.823 |

ΔVT = PET2 − PET1. Panel A pFDR values are corrected across the two ROIs separately within each treatment arm; Panel B pFDR values are corrected across the two ROI interaction tests. Because baseline VT is mathematically contained in ΔVT, these associations should be interpreted cautiously with respect to regression to the mean and measurement variability.

### Supplementary Table 8. Partial correlations between ΔVT and ΔMADRS adjusted for baseline VT.

| Timepoint | Treatment | ROI | n | partial r | 95% CI | p | pFDR |
|---|---|---|---|---|---|---|---|
| Day 8 | Psilocybin | Frontal cortex | 15 | 0.361 | -0.209 to 0.748 | 0.204 | 0.272 |
| Day 8 | Psilocybin | Hippocampus | 15 | 0.371 | -0.199 to 0.753 | 0.192 | 0.272 |
| Day 8 | Niacin | Frontal cortex | 17 | 0.000 | -0.496 to 0.496 | >0.99 | >0.99 |
| Day 8 | Niacin | Hippocampus | 17 | -0.052 | -0.534 to 0.456 | 0.849 | 0.970 |
| PET2 (Day 15) | Psilocybin | Frontal cortex | 15 | 0.292 | -0.282 to 0.712 | 0.311 | 0.323 |
| PET2 (Day 15) | Psilocybin | Hippocampus | 15 | 0.285 | -0.289 to 0.709 | 0.323 | 0.323 |
| PET2 (Day 15) | Niacin | Frontal cortex | 17 | 0.073 | -0.439 to 0.549 | 0.789 | 0.970 |
| PET2 (Day 15) | Niacin | Hippocampus | 17 | 0.053 | -0.454 to 0.535 | 0.845 | 0.970 |
| Day 42 | Psilocybin | Frontal cortex | 15 | 0.605 | 0.110 to 0.860 | 0.022 | 0.107 |
| Day 42 | Psilocybin | Hippocampus | 15 | 0.589 | 0.085 to 0.853 | 0.027 | 0.107 |
| Day 42 | Niacin | Frontal cortex | 16 | 0.158 | -0.386 to 0.620 | 0.575 | 0.970 |
| Day 42 | Niacin | Hippocampus | 16 | 0.168 | -0.377 to 0.626 | 0.550 | 0.970 |
| Day 365 | Psilocybin | Frontal cortex | 10 | 0.527 | -0.211 to 0.882 | 0.145 | 0.272 |
| Day 365 | Psilocybin | Hippocampus | 10 | 0.586 | -0.127 to 0.900 | 0.097 | 0.259 |
| Day 365 | Niacin | Frontal cortex | 9 | -0.405 | -0.863 to 0.419 | 0.319 | 0.970 |
| Day 365 | Niacin | Hippocampus | 9 | -0.457 | -0.879 to 0.365 | 0.255 | 0.970 |

Partial Pearson correlations residualize both ΔVT and ΔMADRS for the corresponding regional baseline VT. pFDR is Benjamini–Hochberg corrected across the eight tests (2 ROIs × 4 timepoints) separately within each treatment arm. Day 42 and Day 365 use censored outcomes.

### Supplementary Table 9. Adjusted within-arm associations between ΔVT and ΔMADRS.

| Timepoint | Treatment | ROI | n | βΔVT | 95% CI | p | pFDR | $R^2$ |
|---|---|---|---|---|---|---|---|---|
| Day 8 | Psilocybin | Frontal cortex | 15 | 0.463 | -0.470 to 1.396 | 0.298 | 0.397 | 0.509 |
| Day 8 | Psilocybin | Hippocampus | 15 | 0.671 | -0.639 to 1.981 | 0.283 | 0.397 | 0.489 |
| Day 8 | Niacin | Frontal cortex | 17 | 0.624 | -3.079 to 4.326 | 0.722 | 0.722 | 0.109 |
| Day 8 | Niacin | Hippocampus | 17 | 1.034 | -4.096 to 6.164 | 0.670 | 0.722 | 0.108 |
| PET2 (Day 15) | Psilocybin | Frontal cortex | 15 | 0.320 | -0.694 to 1.333 | 0.502 | 0.555 | 0.553 |
| PET2 (Day 15) | Psilocybin | Hippocampus | 15 | 0.382 | -1.000 to 1.764 | 0.555 | 0.555 | 0.562 |
| PET2 (Day 15) | Niacin | Frontal cortex | 17 | 1.280 | -1.904 to 4.464 | 0.401 | 0.601 | 0.234 |
| PET2 (Day 15) | Niacin | Hippocampus | 17 | 2.592 | -1.520 to 6.704 | 0.196 | 0.601 | 0.334 |
| Day 42 | Psilocybin | Frontal cortex | 15 | 1.105 | 0.093 to 2.118 | 0.035 | 0.156 | 0.531 |
| Day 42 | Psilocybin | Hippocampus | 15 | 1.472 | 0.088 to 2.857 | 0.039 | 0.156 | 0.537 |
| Day 42 | Niacin | Frontal cortex | 16 | 1.117 | -2.005 to 4.238 | 0.451 | 0.601 | 0.193 |
| Day 42 | Niacin | Hippocampus | 16 | 2.811 | -1.865 to 7.486 | 0.215 | 0.601 | 0.315 |
| Day 365 | Psilocybin | Frontal cortex | 10 | 0.858 | -0.359 to 2.076 | 0.135 | 0.271 | 0.746 |
| Day 365 | Psilocybin | Hippocampus | 10 | 1.293 | -0.275 to 2.860 | 0.090 | 0.240 | 0.782 |
| Day 365 | Niacin | Frontal cortex | 9 | -1.999 | -7.552 to 3.554 | 0.397 | 0.601 | 0.195 |
| Day 365 | Niacin | Hippocampus | 9 | -5.253 | -16.935 to 6.429 | 0.300 | 0.601 | 0.319 |

Models: ΔMADRS ~ baseline MADRS + baseline VT + ΔVT. βΔVT is the coefficient for ΔVT after adjustment for baseline MADRS and the corresponding regional baseline VT. pFDR is corrected across the eight tests separately within each treatment arm. Day 42 and Day 365 use censored outcomes.

### Supplementary Table 10. Fully interacted treatment-robustness models for the association between ΔVT and ΔMADRS.

| Timepoint | ROI | n total | n psilocybin | n niacin | βinteraction | 95% CI | p | pFDR | $R^2$ |
|---|---|---|---|---|---|---|---|---|---|
| Day 8 | Frontal cortex | 32 | 15 | 17 | -0.160 | -3.367 to 3.046 | 0.919 | >0.99 | 0.394 |
| Day 8 | Hippocampus | 32 | 15 | 17 | -0.363 | -4.821 to 4.095 | 0.868 | >0.99 | 0.388 |
| PET2 (Day 15) | Frontal cortex | 32 | 15 | 17 | -0.961 | -3.893 to 1.972 | 0.505 | 0.849 | 0.615 |
| PET2 (Day 15) | Hippocampus | 32 | 15 | 17 | -2.210 | -6.068 to 1.648 | 0.249 | 0.663 | 0.651 |
| Day 42 | Frontal cortex | 31 | 15 | 16 | -0.011 | -2.894 to 2.871 | >0.99 | >0.99 | 0.539 |
| Day 42 | Hippocampus | 31 | 15 | 16 | -1.339 | -5.685 to 3.008 | 0.530 | 0.849 | 0.587 |
| Day 365 | Frontal cortex | 19 | 10 | 9 | 2.857 | -1.683 to 7.398 | 0.193 | 0.663 | 0.631 |
| Day 365 | Hippocampus | 19 | 10 | 9 | 6.546 | -2.832 to 15.923 | 0.153 | 0.663 | 0.686 |

Models: ΔMADRS ~ treatment × (baseline MADRS + baseline VT + ΔVT). βinteraction is the ΔVT × treatment coefficient, allowing the baseline-MADRS and baseline-VT slopes to differ by treatment. Niacin is the reference group. pFDR is corrected across the eight interaction tests. Day 42 and Day 365 use censored outcomes.

**Supplementary Table 11.** Exploratory associations between PET measures and psychological questionnaire outcomes.

| Treatment | Imaging outcome | Questionnaire predictor | n | Spearman's ρ | p | pFDR |
|---|---|---|---|---|---|---|
| Niacin | dBPND_HIP | MEQ_sum | 17 | 0.13 | 0.632 | 0.963 |
| Niacin | dBPND_FC | MEQ_sum | 17 | 0.12 | 0.639 | 0.963 |
| Niacin | dVT_HIP | MEQ_sum | 17 | 0.02 | 0.952 | 0.963 |
| Niacin | dVT_FC | MEQ_sum | 17 | -0.01 | 0.963 | 0.963 |
| Psilocybin | dVT_HIP | MEQ_sum | 15 | -0.32 | 0.237 | 0.752 |
| Psilocybin | dVT_FC | MEQ_sum | 15 | -0.25 | 0.376 | 0.752 |
| Psilocybin | dBPND_HIP | MEQ_sum | 17 | -0.12 | 0.653 | 0.870 |
| Psilocybin | dBPND_FC | MEQ_sum | 17 | -0.04 | 0.870 | 0.870 |
| Niacin | dBPND_HIP | SPIR | 17 | 0.08 | 0.753 | 0.996 |
| Niacin | dVT_HIP | SPIR | 17 | -0.07 | 0.803 | 0.996 |
| Niacin | dVT_FC | SPIR | 17 | -0.04 | 0.867 | 0.996 |
| Niacin | dBPND_FC | SPIR | 17 | -0.00 | 0.996 | 0.996 |
| Psilocybin | dBPND_FC | SPIR | 17 | 0.10 | 0.690 | 0.938 |
| Psilocybin | dVT_HIP | SPIR | 15 | 0.08 | 0.786 | 0.938 |
| Psilocybin | dBPND_HIP | SPIR | 17 | -0.04 | 0.877 | 0.938 |
| Psilocybin | dVT_FC | SPIR | 15 | 0.02 | 0.938 | 0.938 |
| Niacin | dBPND_HIP | dAAQ_D15 | 14 | 0.30 | 0.293 | 0.776 |
| Niacin | dBPND_FC | dAAQ_D15 | 14 | 0.23 | 0.434 | 0.776 |
| Niacin | dVT_FC | dAAQ_D15 | 14 | 0.16 | 0.582 | 0.776 |
| Niacin | dVT_HIP | dAAQ_D15 | 14 | 0.08 | 0.776 | 0.776 |
| Psilocybin | dBPND_FC | dAAQ_D15 | 17 | 0.15 | 0.568 | 0.964 |
| Psilocybin | dBPND_HIP | dAAQ_D15 | 17 | -0.04 | 0.869 | 0.964 |
| Psilocybin | dVT_FC | dAAQ_D15 | 15 | 0.02 | 0.939 | 0.964 |
| Psilocybin | dVT_HIP | dAAQ_D15 | 15 | -0.01 | 0.964 | 0.964 |
| Niacin | dBPND_HIP | MEQ_ineff_mean | 17 | 0.37 | 0.144 | 0.992 |
| Niacin | dBPND_FC | MEQ_positive_mean | 17 | 0.29 | 0.256 | 0.992 |
| Niacin | dBPND_FC | MEQ_ineff_mean | 17 | 0.28 | 0.269 | 0.992 |
| Niacin | dBPND_HIP | MEQ_positive_mean | 17 | 0.25 | 0.324 | 0.992 |
| Niacin | dBPND_HIP | MEQ_timespace_mean | 17 | 0.20 | 0.452 | 0.992 |
| Niacin | dBPND_FC | MEQ_timespace_mean | 17 | 0.14 | 0.600 | 0.992 |
| Niacin | dVT_FC | MEQ_timespace_mean | 17 | -0.10 | 0.707 | 0.992 |
| Niacin | dVT_FC | MEQ_ineff_mean | 17 | 0.09 | 0.726 | 0.992 |
| Niacin | dVT_HIP | MEQ_ineff_mean | 17 | 0.07 | 0.788 | 0.992 |
| Niacin | dVT_HIP | MEQ_timespace_mean | 17 | -0.05 | 0.853 | 0.992 |
| Niacin | dVT_FC | MEQ_mystical_mean | 17 | -0.05 | 0.857 | 0.992 |
| Niacin | dVT_HIP | MEQ_mystical_mean | 17 | -0.03 | 0.910 | 0.992 |
| Niacin | dVT_HIP | MEQ_positive_mean | 17 | 0.01 | 0.962 | 0.992 |
| Niacin | dBPND_FC | MEQ_mystical_mean | 17 | -0.01 | 0.962 | 0.992 |
| Niacin | dVT_FC | MEQ_positive_mean | 17 | 0.01 | 0.977 | 0.992 |
| Niacin | dBPND_HIP | MEQ_mystical_mean | 17 | -0.00 | 0.992 | 0.992 |
| Psilocybin | dVT_HIP | MEQ_mystical_mean | 15 | -0.39 | 0.147 | 0.605 |
| Psilocybin | dVT_FC | MEQ_ineff_mean | 15 | -0.38 | 0.164 | 0.605 |
| Psilocybin | dVT_HIP | MEQ_timespace_mean | 15 | -0.35 | 0.199 | 0.605 |
| Psilocybin | dVT_HIP | MEQ_ineff_mean | 15 | -0.35 | 0.206 | 0.605 |
| Psilocybin | dBPND_FC | MEQ_ineff_mean | 17 | -0.30 | 0.250 | 0.605 |
| Psilocybin | dBPND_HIP | MEQ_ineff_mean | 17 | -0.29 | 0.259 | 0.605 |
| Psilocybin | dVT_FC | MEQ_mystical_mean | 15 | -0.31 | 0.265 | 0.605 |
| Psilocybin | dBPND_HIP | MEQ_timespace_mean | 17 | -0.27 | 0.303 | 0.605 |
| Psilocybin | dVT_FC | MEQ_timespace_mean | 15 | -0.24 | 0.384 | 0.682 |
| Psilocybin | dVT_HIP | MEQ_positive_mean | 15 | -0.21 | 0.450 | 0.720 |
| Psilocybin | dBPND_FC | MEQ_timespace_mean | 17 | -0.17 | 0.517 | 0.752 |
| Psilocybin | dVT_FC | MEQ_positive_mean | 15 | -0.14 | 0.615 | 0.804 |
| Psilocybin | dBPND_HIP | MEQ_mystical_mean | 17 | -0.12 | 0.653 | 0.804 |
| Psilocybin | dBPND_HIP | MEQ_positive_mean | 17 | 0.09 | 0.718 | 0.820 |
| Psilocybin | dBPND_FC | MEQ_mystical_mean | 17 | -0.06 | 0.823 | 0.837 |
| Psilocybin | dBPND_FC | MEQ_positive_mean | 17 | 0.05 | 0.837 | 0.837 |
| Niacin | dBPND_FC | CEQ_sum | 17 | -0.08 | 0.754 | 0.985 |
| Niacin | dBPND_HIP | CEQ_sum | 17 | 0.05 | 0.844 | 0.985 |
| Niacin | dVT_FC | CEQ_sum | 17 | -0.03 | 0.907 | 0.985 |
| Niacin | dVT_HIP | CEQ_sum | 17 | -0.00 | 0.985 | 0.985 |
| Psilocybin | dBPND_HIP | CEQ_sum | 17 | -0.40 | 0.111 | 0.398 |
| Psilocybin | dBPND_FC | CEQ_sum | 17 | -0.29 | 0.256 | 0.398 |
| Psilocybin | dVT_HIP | CEQ_sum | 15 | -0.29 | 0.298 | 0.398 |
| Psilocybin | dVT_FC | CEQ_sum | 15 | -0.18 | 0.528 | 0.528 |

| Treatment | Imaging outcome | Questionnaire predictor | n | Spearman's ρ | p | pFDR |
|---|---|---|---|---|---|---|
| Niacin | dVT_HIP | EBI_mean | 17 | -0.16 | 0.549 | 0.943 |
| Niacin | dBPND_HIP | EBI_mean | 17 | -0.12 | 0.641 | 0.943 |
| Niacin | dVT_FC | EBI_mean | 17 | -0.10 | 0.707 | 0.943 |
| Niacin | dBPND_FC | EBI_mean | 17 | -0.02 | 0.944 | 0.944 |
| Psilocybin | dVT_HIP | EBI_mean | 15 | -0.32 | 0.250 | 0.528 |
| Psilocybin | dVT_FC | EBI_mean | 15 | -0.31 | 0.264 | 0.528 |
| Psilocybin | dBPND_HIP | EBI_mean | 17 | -0.12 | 0.656 | 0.874 |
| Psilocybin | dBPND_FC | EBI_mean | 17 | 0.02 | 0.944 | 0.944 |
| Niacin | dVT_FC | LEQ_delta_365 | 15 | -0.55 | 0.034 | 0.234 |
| Niacin | dBPND_FC | LEQ_delta_42 | 16 | -0.50 | 0.048 | 0.234 |
| Niacin | dBPND_FC | LEQ_delta_365 | 15 | -0.48 | 0.069 | 0.234 |
| Niacin | dBPND_HIP | LEQ_delta_42 | 16 | -0.45 | 0.078 | 0.234 |
| Niacin | dVT_HIP | LEQ_delta_365 | 15 | -0.39 | 0.155 | 0.372 |
| Niacin | dBPND_HIP | LEQ_delta_365 | 15 | -0.20 | 0.466 | 0.876 |
| Niacin | dBPND_HIP | LEQ_delta_15 | 13 | 0.18 | 0.566 | 0.876 |
| Niacin | dBPND_FC | LEQ_delta_15 | 13 | -0.16 | 0.603 | 0.876 |
| Niacin | dVT_HIP | LEQ_delta_42 | 16 | -0.10 | 0.716 | 0.876 |
| Niacin | dVT_FC | LEQ_delta_42 | 16 | -0.08 | 0.766 | 0.876 |
| Niacin | dVT_FC | LEQ_delta_15 | 13 | -0.08 | 0.803 | 0.876 |
| Niacin | dVT_HIP | LEQ_delta_15 | 13 | -0.01 | 0.972 | 0.972 |
| Psilocybin | dVT_FC | LEQ_delta_365 | 15 | -0.74 | 0.002 | 0.019 |
| Psilocybin | dVT_HIP | LEQ_delta_365 | 15 | -0.70 | 0.004 | 0.021 |
| Psilocybin | dVT_FC | LEQ_delta_15 | 15 | -0.60 | 0.017 | 0.070 |
| Psilocybin | dVT_HIP | LEQ_delta_15 | 15 | -0.58 | 0.024 | 0.073 |
| Psilocybin | dBPND_FC | LEQ_delta_15 | 17 | -0.38 | 0.134 | 0.306 |
| Psilocybin | dVT_FC | LEQ_delta_42 | 13 | -0.41 | 0.168 | 0.306 |
| Psilocybin | dBPND_FC | LEQ_delta_365 | 17 | -0.32 | 0.216 | 0.306 |
| Psilocybin | dBPND_HIP | LEQ_delta_365 | 17 | -0.30 | 0.235 | 0.306 |
| Psilocybin | dBPND_HIP | LEQ_delta_15 | 17 | -0.29 | 0.254 | 0.306 |
| Psilocybin | dVT_HIP | LEQ_delta_42 | 13 | -0.34 | 0.255 | 0.306 |
| Psilocybin | dBPND_FC | LEQ_delta_42 | 15 | -0.22 | 0.420 | 0.458 |
| Psilocybin | dBPND_HIP | LEQ_delta_42 | 15 | -0.08 | 0.781 | 0.781 |
| Niacin | dVT_FC | MLQ_mean_delta15 | 15 | -0.44 | 0.099 | 0.670 |
| Niacin | dVT_HIP | MLQ_mean_delta15 | 15 | -0.41 | 0.133 | 0.670 |
| Niacin | dBPND_HIP | MLQ_mean_delta365 | 15 | -0.29 | 0.289 | 0.670 |
| Niacin | dBPND_FC | MLQ_mean_delta365 | 15 | -0.29 | 0.298 | 0.670 |
| Niacin | dVT_HIP | MLQ_mean_delta42 | 17 | -0.26 | 0.320 | 0.670 |
| Niacin | dVT_FC | MLQ_mean_delta42 | 17 | -0.25 | 0.335 | 0.670 |
| Niacin | dVT_FC | MLQ_mean_delta365 | 15 | -0.15 | 0.602 | 0.924 |
| Niacin | dBPND_HIP | MLQ_mean_delta42 | 17 | 0.12 | 0.649 | 0.924 |
| Niacin | dBPND_FC | MLQ_mean_delta42 | 17 | 0.09 | 0.725 | 0.924 |
| Niacin | dBPND_FC | MLQ_mean_delta15 | 15 | -0.08 | 0.785 | 0.924 |
| Niacin | dVT_HIP | MLQ_mean_delta365 | 15 | -0.05 | 0.849 | 0.924 |
| Niacin | dBPND_HIP | MLQ_mean_delta15 | 15 | 0.03 | 0.924 | 0.924 |
| Psilocybin | dVT_HIP | MLQ_mean_delta15 | 15 | -0.19 | 0.507 | 0.994 |
| Psilocybin | dBPND_FC | MLQ_mean_delta15 | 17 | -0.12 | 0.639 | 0.994 |
| Psilocybin | dBPND_HIP | MLQ_mean_delta365 | 16 | -0.13 | 0.641 | 0.994 |
| Psilocybin | dVT_FC | MLQ_mean_delta365 | 14 | 0.13 | 0.648 | 0.994 |
| Psilocybin | dBPND_FC | MLQ_mean_delta365 | 16 | -0.10 | 0.704 | 0.994 |
| Psilocybin | dVT_FC | MLQ_mean_delta15 | 15 | -0.11 | 0.708 | 0.994 |
| Psilocybin | dBPND_HIP | MLQ_mean_delta15 | 17 | -0.09 | 0.740 | 0.994 |
| Psilocybin | dVT_HIP | MLQ_mean_delta42 | 13 | -0.06 | 0.858 | 0.994 |
| Psilocybin | dBPND_FC | MLQ_mean_delta42 | 15 | -0.04 | 0.879 | 0.994 |
| Psilocybin | dBPND_HIP | MLQ_mean_delta42 | 15 | 0.02 | 0.950 | 0.994 |
| Psilocybin | dVT_FC | MLQ_mean_delta42 | 13 | -0.00 | 0.993 | 0.994 |
| Psilocybin | dVT_HIP | MLQ_mean_delta365 | 14 | 0.00 | 0.994 | 0.994 |
| Niacin | dBPND_HIP | SWLS_mean_delta15 | 15 | 0.20 | 0.472 | 0.949 |
| Niacin | dBPND_HIP | SWLS_mean_delta365 | 15 | 0.20 | 0.475 | 0.949 |
| Niacin | dVT_HIP | SWLS_mean_delta365 | 15 | -0.10 | 0.723 | 0.949 |
| Niacin | dVT_FC | SWLS_mean_delta365 | 15 | -0.10 | 0.723 | 0.949 |
| Niacin | dVT_HIP | SWLS_mean_delta42 | 17 | -0.08 | 0.755 | 0.949 |
| Niacin | dVT_FC | SWLS_mean_delta15 | 15 | -0.09 | 0.755 | 0.949 |
| Niacin | dBPND_HIP | SWLS_mean_delta42 | 17 | 0.07 | 0.777 | 0.949 |
| Niacin | dVT_HIP | SWLS_mean_delta15 | 15 | -0.04 | 0.899 | 0.949 |
| Niacin | dVT_FC | SWLS_mean_delta42 | 17 | -0.03 | 0.902 | 0.949 |
| Niacin | dBPND_FC | SWLS_mean_delta42 | 17 | 0.02 | 0.936 | 0.949 |
| Niacin | dBPND_FC | SWLS_mean_delta365 | 15 | 0.02 | 0.940 | 0.949 |

| Treatment | Imaging outcome | Questionnaire predictor | n | Spearman's ρ | p | pFDR |
|---|---|---|---|---|---|---|
| Niacin | dBPND_FC | SWLS_mean_delta15 | 15 | 0.02 | 0.949 | 0.949 |
| Psilocybin | dVT_HIP | SWLS_mean_delta365 | 14 | -0.51 | 0.065 | 0.417 |
| Psilocybin | dVT_FC | SWLS_mean_delta365 | 14 | -0.45 | 0.107 | 0.417 |
| Psilocybin | dVT_FC | SWLS_mean_delta15 | 15 | -0.38 | 0.164 | 0.417 |
| Psilocybin | dBPND_FC | SWLS_mean_delta15 | 17 | -0.33 | 0.196 | 0.417 |
| Psilocybin | dVT_HIP | SWLS_mean_delta15 | 15 | -0.35 | 0.201 | 0.417 |
| Psilocybin | dBPND_HIP | SWLS_mean_delta15 | 17 | -0.29 | 0.260 | 0.417 |
| Psilocybin | dBPND_HIP | SWLS_mean_delta365 | 16 | -0.29 | 0.268 | 0.417 |
| Psilocybin | dBPND_FC | SWLS_mean_delta365 | 16 | -0.28 | 0.289 | 0.417 |
| Psilocybin | dVT_FC | SWLS_mean_delta42 | 13 | -0.27 | 0.365 | 0.417 |
| Psilocybin | dBPND_FC | SWLS_mean_delta42 | 15 | -0.25 | 0.367 | 0.417 |
| Psilocybin | dBPND_HIP | SWLS_mean_delta42 | 15 | -0.24 | 0.385 | 0.417 |
| Psilocybin | dVT_HIP | SWLS_mean_delta42 | 13 | -0.25 | 0.417 | 0.417 |

Note. Spearman rank correlations (ρ) between changes in [$^{11}$C]UCB-J binding (PET2 − PET1) and psychological questionnaire measures, calculated separately within each treatment group. Sample size (n) denotes participants with complete data for both the PET measure and the corresponding questionnaire. FDR-adjusted p values were calculated using the Benjamini–Hochberg procedure separately within each treatment group and questionnaire family, across the PET measures and questionnaire variables included in that family. ΔPET = PET2 − PET1; FC, frontal cortex; HIP, hippocampus; VT, total distribution volume; BPND, non-displaceable binding potential.

### Supplementary Table 12. Exploratory ANCOVA analyses applying the primary model to an extended set of anatomically defined VT regions of interest.

| Model | Measure | Region | β | SE | 95% CI | p | N | R² | Adj. R² |
|---|---|---|---|---|---|---|---|---|---|
| 1 | VT | HIP | 0.053 | 0.71 | -1.39 to 1.50 | 0.940 | 32 | 0.24 | 0.19 |
| 1 | VT | FC | -0.49 | 0.94 | -2.42 to 1.44 | 0.607 | 32 | 0.12 | 0.05 |
| 1 | VT | GM | -0.41 | 0.83 | -2.10 to 1.29 | 0.626 | 32 | 0.14 | 0.08 |
| 1 | VT | AMY | -0.053 | 1.03 | -2.16 to 2.06 | 0.959 | 32 | 0.17 | 0.11 |
| 1 | VT | THA | 0.014 | 0.69 | -1.40 to 1.42 | 0.984 | 32 | 0.30 | 0.25 |
| 1 | VT | PUT | -0.55 | 0.98 | -2.56 to 1.45 | 0.577 | 32 | 0.21 | 0.15 |
| 1 | VT | CAU | -0.29 | 1.03 | -2.40 to 1.82 | 0.780 | 32 | 0.22 | 0.17 |
| 1 | VT | INS | -0.46 | 0.97 | -2.44 to 1.52 | 0.636 | 32 | 0.12 | 0.06 |
| 1 | VT | BST | 0.028 | 0.29 | -0.57 to 0.62 | 0.925 | 32 | 0.34 | 0.30 |
| 1 | VT | cACC | -1.04 | 0.93 | -2.95 to 0.87 | 0.274 | 32 | 0.23 | 0.18 |
| 1 | VT | rACC | -0.80 | 0.97 | -2.79 to 1.19 | 0.417 | 32 | 0.22 | 0.16 |
| 1 | VT | ACC | -0.91 | 0.94 | -2.83 to 1.00 | 0.338 | 32 | 0.21 | 0.16 |
| 1 | VT | rMFG | -0.38 | 0.97 | -2.38 to 1.61 | 0.697 | 32 | 0.11 | 0.04 |
| 1 | VT | mOFC | -0.34 | 1.05 | -2.48 to 1.80 | 0.745 | 32 | 0.08 | 0.01 |
| 1 | VT | CSO | -0.066 | 0.14 | -0.35 to 0.22 | 0.639 | 32 | 0.45 | 0.41 |
| 2 | VT | HIP | 0.12 | 0.71 | -1.33 to 1.57 | 0.867 | 32 | 0.26 | 0.18 |
| 2 | VT | FC | -0.38 | 0.94 | -2.30 to 1.53 | 0.686 | 32 | 0.17 | 0.08 |
| 2 | VT | GM | -0.33 | 0.83 | -2.03 to 1.37 | 0.695 | 32 | 0.17 | 0.08 |
| 2 | VT | AMY | 0.050 | 1.03 | -2.06 to 2.16 | 0.961 | 32 | 0.20 | 0.12 |
| 2 | VT | THA | 0.053 | 0.70 | -1.38 to 1.49 | 0.941 | 32 | 0.31 | 0.23 |
| 2 | VT | PUT | -0.47 | 0.99 | -2.48 to 1.55 | 0.640 | 32 | 0.23 | 0.15 |
| 2 | VT | CAU | -0.17 | 1.02 | -2.26 to 1.92 | 0.867 | 32 | 0.27 | 0.19 |
| 2 | VT | INS | -0.34 | 0.95 | -2.30 to 1.61 | 0.722 | 32 | 0.18 | 0.09 |
| 2 | VT | BST | 0.057 | 0.29 | -0.54 to 0.65 | 0.845 | 32 | 0.37 | 0.30 |
| 2 | VT | cACC | -0.94 | 0.93 | -2.84 to 0.96 | 0.318 | 32 | 0.27 | 0.19 |
| 2 | VT | rACC | -0.72 | 0.98 | -2.71 to 1.28 | 0.469 | 32 | 0.24 | 0.16 |
| 2 | VT | ACC | -0.83 | 0.94 | -2.74 to 1.09 | 0.383 | 32 | 0.25 | 0.16 |
| 2 | VT | rMFG | -0.26 | 0.96 | -2.24 to 1.71 | 0.786 | 32 | 0.16 | 0.07 |
| 2 | VT | mOFC | -0.19 | 1.02 | -2.27 to 1.89 | 0.857 | 32 | 0.17 | 0.08 |
| 2 | VT | CSO | -0.053 | 0.14 | -0.34 to 0.23 | 0.704 | 32 | 0.47 | 0.41 |
| 3 | VT | HIP | 0.54 | 0.75 | -1.01 to 2.10 | 0.474 | 28 | 0.45 | 0.26 |
| 3 | VT | FC | 0.29 | 0.96 | -1.71 to 2.29 | 0.767 | 28 | 0.38 | 0.17 |
| 3 | VT | GM | 0.27 | 0.83 | -1.46 to 2.00 | 0.751 | 28 | 0.42 | 0.22 |
| 3 | VT | AMY | 0.52 | 1.20 | -1.97 to 3.02 | 0.667 | 28 | 0.31 | 0.07 |
| 3 | VT | THA | 0.64 | 0.67 | -0.75 to 2.02 | 0.349 | 28 | 0.58 | 0.43 |
| 3 | VT | PUT | 0.34 | 1.01 | -1.76 to 2.44 | 0.738 | 28 | 0.40 | 0.19 |
| 3 | VT | CAU | 0.53 | 1.01 | -1.59 to 2.64 | 0.608 | 28 | 0.47 | 0.29 |
| 3 | VT | INS | 0.24 | 1.02 | -1.88 to 2.37 | 0.814 | 28 | 0.36 | 0.14 |
| 3 | VT | BST | 0.33 | 0.29 | -0.28 to 0.93 | 0.272 | 28 | 0.59 | 0.45 |

Psilocybin and synaptic density in depression

| Model | Measure | Region | β | SE | 95% CI | p | N | R² | Adj. R² |
|---|---|---|---|---|---|---|---|---|---|
| 3 | VT | cACC | -0.58 | 0.97 | -2.61 to 1.45 | 0.557 | 28 | 0.49 | 0.32 |
| 3 | VT | rACC | 0.34 | 1.02 | -1.79 to 2.47 | 0.743 | 28 | 0.47 | 0.29 |
| 3 | VT | ACC | -0.082 | 0.98 | -2.13 to 1.96 | 0.934 | 28 | 0.47 | 0.29 |
| 3 | VT | rMFG | 0.38 | 1.00 | -1.70 to 2.45 | 0.709 | 28 | 0.36 | 0.14 |
| 3 | VT | mOFC | 0.60 | 1.09 | -1.68 to 2.88 | 0.591 | 28 | 0.38 | 0.17 |
| 3 | VT | CSO | -0.008 | 0.15 | -0.33 to 0.31 | 0.957 | 28 | 0.55 | 0.39 |

The same ANCOVA framework as the primary analysis was applied to additional anatomically defined ROIs. These analyses are hypothesis-generating.

### Supplementary Table 13. Exploratory ANCOVA analyses applying the primary model to an extended set of anatomically defined BPND regions of interest.

| Model | Measure | Region | β | SE | 95% CI | p | N | $R^2$ | Adj. $R^2$ |
|---|---|---|---|---|---|---|---|---|---|
| 1 | BPND | HIP | 0.013 | 0.073 | -0.14 to 0.16 | 0.858 | 34 | 0.38 | 0.34 |
| 1 | BPND | FC | 0.010 | 0.11 | -0.22 to 0.24 | 0.931 | 34 | 0.36 | 0.32 |
| 1 | BPND | GM | 0.004 | 0.094 | -0.19 to 0.20 | 0.964 | 34 | 0.41 | 0.37 |
| 1 | BPND | AMY | 0.020 | 0.093 | -0.17 to 0.21 | 0.835 | 34 | 0.33 | 0.29 |
| 1 | BPND | THA | 0.023 | 0.074 | -0.13 to 0.17 | 0.754 | 34 | 0.41 | 0.38 |
| 1 | BPND | PUT | 0.018 | 0.13 | -0.24 to 0.27 | 0.889 | 34 | 0.42 | 0.38 |
| 1 | BPND | CAU | 0.023 | 0.11 | -0.21 to 0.25 | 0.838 | 34 | 0.41 | 0.37 |
| 1 | BPND | INS | 0.021 | 0.11 | -0.20 to 0.25 | 0.847 | 34 | 0.50 | 0.47 |
| 1 | BPND | BST | 0.025 | 0.036 | -0.049 to 0.098 | 0.502 | 34 | 0.29 | 0.24 |
| 1 | BPND | cACC | -0.031 | 0.11 | -0.27 to 0.20 | 0.786 | 34 | 0.56 | 0.53 |
| 1 | BPND | rACC | 0.017 | 0.12 | -0.22 to 0.26 | 0.884 | 34 | 0.54 | 0.51 |
| 1 | BPND | ACC | -0.004 | 0.11 | -0.23 to 0.23 | 0.971 | 34 | 0.55 | 0.52 |
| 1 | BPND | rMFG | 0.057 | 0.12 | -0.20 to 0.31 | 0.648 | 34 | 0.34 | 0.30 |
| 1 | BPND | mOFC | 0.044 | 0.12 | -0.21 to 0.30 | 0.726 | 34 | 0.38 | 0.34 |
| 2 | BPND | HIP | 0.013 | 0.075 | -0.14 to 0.17 | 0.863 | 34 | 0.38 | 0.32 |
| 2 | BPND | FC | 0.013 | 0.12 | -0.22 to 0.25 | 0.914 | 34 | 0.37 | 0.30 |
| 2 | BPND | GM | 0.004 | 0.096 | -0.19 to 0.20 | 0.967 | 34 | 0.41 | 0.35 |
| 2 | BPND | AMY | 0.020 | 0.095 | -0.17 to 0.21 | 0.838 | 34 | 0.33 | 0.26 |
| 2 | BPND | THA | 0.022 | 0.075 | -0.13 to 0.18 | 0.769 | 34 | 0.42 | 0.36 |
| 2 | BPND | PUT | 0.016 | 0.13 | -0.25 to 0.28 | 0.901 | 34 | 0.42 | 0.36 |
| 2 | BPND | CAU | 0.025 | 0.11 | -0.21 to 0.26 | 0.825 | 34 | 0.41 | 0.35 |
| 2 | BPND | INS | 0.024 | 0.11 | -0.20 to 0.25 | 0.829 | 34 | 0.51 | 0.46 |
| 2 | BPND | BST | 0.027 | 0.036 | -0.047 to 0.10 | 0.468 | 34 | 0.31 | 0.24 |
| 2 | BPND | cACC | -0.029 | 0.12 | -0.27 to 0.21 | 0.803 | 34 | 0.56 | 0.52 |
| 2 | BPND | rACC | 0.017 | 0.12 | -0.23 to 0.26 | 0.889 | 34 | 0.54 | 0.50 |
| 2 | BPND | ACC | -0.004 | 0.11 | -0.24 to 0.23 | 0.974 | 34 | 0.55 | 0.51 |
| 2 | BPND | rMFG | 0.060 | 0.13 | -0.20 to 0.32 | 0.634 | 34 | 0.35 | 0.28 |
| 2 | BPND | mOFC | 0.051 | 0.12 | -0.20 to 0.31 | 0.682 | 34 | 0.40 | 0.34 |
| 3 | BPND | HIP | 0.081 | 0.073 | -0.072 to 0.23 | 0.282 | 29 | 0.64 | 0.52 |
| 3 | BPND | FC | 0.10 | 0.11 | -0.13 to 0.33 | 0.364 | 29 | 0.65 | 0.54 |
| 3 | BPND | GM | 0.079 | 0.091 | -0.11 to 0.27 | 0.395 | 29 | 0.67 | 0.56 |
| 3 | BPND | AMY | 0.11 | 0.095 | -0.083 to 0.31 | 0.243 | 29 | 0.59 | 0.46 |
| 3 | BPND | THA | 0.085 | 0.074 | -0.069 to 0.24 | 0.264 | 29 | 0.64 | 0.52 |
| 3 | BPND | PUT | 0.15 | 0.12 | -0.10 to 0.41 | 0.229 | 29 | 0.66 | 0.54 |
| 3 | BPND | CAU | 0.14 | 0.11 | -0.083 to 0.36 | 0.207 | 29 | 0.68 | 0.57 |
| 3 | BPND | INS | 0.12 | 0.10 | -0.094 to 0.33 | 0.257 | 29 | 0.74 | 0.66 |
| 3 | BPND | BST | 0.047 | 0.043 | -0.042 to 0.14 | 0.282 | 29 | 0.44 | 0.25 |
| 3 | BPND | cACC | 0.050 | 0.12 | -0.19 to 0.29 | 0.674 | 29 | 0.70 | 0.61 |
| 3 | BPND | rACC | 0.18 | 0.11 | -0.057 to 0.42 | 0.128 | 29 | 0.74 | 0.65 |
| 3 | BPND | ACC | 0.12 | 0.11 | -0.11 to 0.35 | 0.281 | 29 | 0.73 | 0.64 |
| 3 | BPND | rMFG | 0.14 | 0.12 | -0.11 to 0.40 | 0.261 | 29 | 0.61 | 0.48 |
| 3 | BPND | mOFC | 0.18 | 0.12 | -0.075 to 0.44 | 0.155 | 29 | 0.62 | 0.50 |

The same ANCOVA framework as the primary analysis was applied to additional anatomically defined ROIs. These analyses are hypothesis-generating.

### Supplementary Table 14. Descriptive SV2A binding in prespecified primary regions of interest.

PET1, PET2, and within-participant changes in $V_t$ and $BP_{nD}$ in frontal cortex and hippocampus by treatment group. Values are unadjusted means (SD), with Δ = PET2 − PET1. Of the 35 participants, 34 completed both PET examinations (17 psilocybin, 17 niacin). $V_t$ could not be estimated for two participants in the psilocybin group due to incomplete arterial blood sampling. Sample sizes reflect participants with complete paired measurements. Adjusted between-group comparisons are reported in Supplementary Table 1.

| Measure | ROI | Treatment | n | PET1 mean (SD) | PET2 mean (SD) | Δ mean (SD) |
|---|---|---|---|---|---|---|
| VT | Frontal cortex | Psilocybin | 15 | 22.86 (3.20) | 22.25 (3.66) | -0.61 (4.09) |
| VT | Frontal cortex | Niacin | 17 | 23.34 (2.64) | 22.88 (1.59) | -0.45 (2.42) |
| VT | Hippocampus | Psilocybin | 15 | 16.18 (2.55) | 16.00 (2.95) | -0.19 (2.79) |
| VT | Hippocampus | Niacin | 17 | 16.28 (2.11) | 15.99 (1.34) | -0.29 (1.79) |
| BPND | Frontal cortex | Psilocybin | 17 | 3.93 (0.48) | 3.87 (0.45) | -0.06 (0.43) |
| BPND | Frontal cortex | Niacin | 17 | 3.86 (0.33) | 3.82 (0.36) | -0.04 (0.28) |
| BPND | Hippocampus | Psilocybin | 17 | 2.34 (0.30) | 2.31 (0.32) | -0.03 (0.26) |
| BPND | Hippocampus | Niacin | 17 | 2.31 (0.21) | 2.28 (0.20) | -0.03 (0.20) |

Values are unadjusted mean (SD). Δ = PET2 − PET1, calculated within participants. n indicates participants with both PET1 and PET2 values for the respective measure and ROI.

### Supplementary Table 15. Descriptive SV2A binding in exploratory regions of interest.

Values are unadjusted means (SD). Δ = PET2 − PET1, calculated within participants. n indicates participants with complete paired measurements. Abbreviations: GM, global grey matter; AMY, amygdala; THA, thalamus; PUT, putamen; CAU, caudate; INS, insula; BST, bed nucleus of the stria terminalis; cACC, caudal anterior cingulate cortex; rACC, rostral anterior cingulate cortex; ACC, anterior cingulate cortex (including cACC and rACC); rMFG, rostral middle frontal gyrus; mOFC, medial orbitofrontal cortex; CSO, centrum semiovale (reference region for $BP_{nD}$). Adjusted between-group comparisons are reported in Supplementary Tables 12–13.

| Measure | ROI | Treatment | n | PET1 mean (SD) | PET2 mean (SD) | Δ mean (SD) |
|---|---|---|---|---|---|---|
| VT | GM | Psilocybin | 15 | 21.17 (2.87) | 20.57 (3.21) | -0.61 (3.58) |
| VT | GM | Niacin | 17 | 21.55 (2.40) | 21.10 (1.50) | -0.45 (2.12) |
| VT | AMY | Psilocybin | 15 | 21.78 (3.98) | 21.71 (4.08) | -0.07 (4.65) |
| VT | AMY | Niacin | 17 | 22.33 (2.85) | 21.97 (1.95) | -0.36 (2.23) |
| VT | THA | Psilocybin | 15 | 16.03 (2.52) | 15.80 (3.06) | -0.23 (2.58) |
| VT | THA | Niacin | 17 | 16.20 (1.89) | 15.88 (1.25) | -0.32 (1.66) |
| VT | PUT | Psilocybin | 15 | 24.99 (3.64) | 24.45 (3.75) | -0.54 (4.34) |
| VT | PUT | Niacin | 17 | 25.85 (3.28) | 25.33 (2.12) | -0.52 (2.44) |
| VT | CAU | Psilocybin | 15 | 22.93 (4.00) | 22.63 (4.26) | -0.31 (4.29) |
| VT | CAU | Niacin | 17 | 23.33 (2.66) | 23.09 (1.90) | -0.23 (2.37) |
| VT | INS | Psilocybin | 15 | 23.19 (3.36) | 22.61 (3.78) | -0.58 (4.18) |
| VT | INS | Niacin | 17 | 23.63 (2.43) | 23.21 (1.61) | -0.42 (2.32) |
| VT | BST | Psilocybin | 15 | 7.84 (1.16) | 7.71 (1.29) | -0.13 (1.12) |
| VT | BST | Niacin | 17 | 8.04 (0.97) | 7.79 (0.61) | -0.25 (0.75) |
| VT | cACC | Psilocybin | 15 | 23.94 (3.57) | 22.96 (3.84) | -0.98 (4.12) |
| VT | cACC | Niacin | 17 | 24.35 (2.65) | 24.17 (1.62) | -0.19 (2.03) |
| VT | rACC | Psilocybin | 15 | 24.72 (3.39) | 24.18 (3.56) | -0.54 (4.22) |
| VT | rACC | Niacin | 17 | 26.19 (3.38) | 25.50 (2.14) | -0.68 (2.51) |
| VT | ACC | Psilocybin | 15 | 24.34 (3.41) | 23.57 (3.65) | -0.77 (4.13) |
| VT | ACC | Niacin | 17 | 25.27 (2.87) | 24.83 (1.80) | -0.44 (2.19) |
| VT | rMFG | Psilocybin | 15 | 23.73 (3.30) | 23.09 (3.81) | -0.64 (4.24) |
| VT | rMFG | Niacin | 17 | 23.99 (2.70) | 23.55 (1.56) | -0.44 (2.50) |
| VT | mOFC | Psilocybin | 15 | 23.31 (3.31) | 22.93 (3.85) | -0.37 (4.29) |
| VT | mOFC | Niacin | 17 | 24.21 (3.07) | 23.49 (1.90) | -0.71 (3.21) |
| VT | CSO | Psilocybin | 15 | 4.17 (0.74) | 4.10 (0.63) | -0.07 (0.55) |
| VT | CSO | Niacin | 17 | 4.26 (0.47) | 4.21 (0.39) | -0.04 (0.39) |
| BPND | GM | Psilocybin | 17 | 3.55 (0.45) | 3.50 (0.38) | -0.05 (0.36) |
| BPND | GM | Niacin | 17 | 3.50 (0.26) | 3.46 (0.31) | -0.04 (0.23) |
| BPND | AMY | Psilocybin | 17 | 2.88 (0.33) | 2.89 (0.35) | 0.01 (0.35) |
| BPND | AMY | Niacin | 17 | 2.91 (0.30) | 2.88 (0.30) | -0.02 (0.23) |

| Measure | ROI | Treatment | n | PET1 mean (SD) | PET2 mean (SD) | Δ mean (SD) |
|---|---|---|---|---|---|---|
| BPND | THA | Psilocybin | 17 | 2.85 (0.27) | 2.83 (0.32) | -0.02 (0.26) |
| BPND | THA | Niacin | 17 | 2.80 (0.18) | 2.77 (0.22) | -0.04 (0.16) |
| BPND | PUT | Psilocybin | 17 | 4.29 (0.54) | 4.24 (0.48) | -0.05 (0.48) |
| BPND | PUT | Niacin | 17 | 4.33 (0.42) | 4.25 (0.47) | -0.09 (0.31) |
| BPND | CAU | Psilocybin | 17 | 3.69 (0.45) | 3.68 (0.44) | -0.01 (0.43) |
| BPND | CAU | Niacin | 17 | 3.70 (0.38) | 3.66 (0.40) | -0.04 (0.26) |
| BPND | INS | Psilocybin | 17 | 3.69 (0.50) | 3.63 (0.47) | -0.06 (0.40) |
| BPND | INS | Niacin | 17 | 3.62 (0.39) | 3.56 (0.42) | -0.06 (0.27) |
| BPND | BST | Psilocybin | 17 | 1.06 (0.21) | 1.07 (0.14) | 0.01 (0.18) |
| BPND | BST | Niacin | 17 | 1.07 (0.08) | 1.05 (0.10) | -0.02 (0.07) |
| BPND | cACC | Psilocybin | 17 | 3.99 (0.61) | 3.90 (0.50) | -0.09 (0.46) |
| BPND | cACC | Niacin | 17 | 3.91 (0.47) | 3.87 (0.49) | -0.03 (0.25) |
| BPND | rACC | Psilocybin | 17 | 3.87 (0.59) | 3.81 (0.50) | -0.06 (0.40) |
| BPND | rACC | Niacin | 17 | 3.96 (0.44) | 3.86 (0.49) | -0.10 (0.34) |
| BPND | ACC | Psilocybin | 17 | 3.93 (0.59) | 3.86 (0.48) | -0.07 (0.42) |
| BPND | ACC | Niacin | 17 | 3.93 (0.44) | 3.86 (0.48) | -0.07 (0.29) |
| BPND | rMFG | Psilocybin | 17 | 4.11 (0.50) | 4.07 (0.48) | -0.04 (0.48) |
| BPND | rMFG | Niacin | 17 | 4.00 (0.36) | 3.95 (0.37) | -0.05 (0.30) |
| BPND | mOFC | Psilocybin | 17 | 3.70 (0.54) | 3.70 (0.45) | -0.01 (0.43) |
| BPND | mOFC | Niacin | 17 | 3.73 (0.41) | 3.67 (0.46) | -0.06 (0.38) |

Values are unadjusted mean (SD). Δ = PET2 − PET1, calculated within participants. n indicates participants with both PET1 and PET2 values for the respective measure and ROI.

**Supplementary Table 16. Deviations from, and additions to, the preregistered analysis plan (AsPredicted #152777).**

The registration is dated 28 November 2023.

| Item | As registered | As reported in the manuscript |
|---|---|---|
| Timing | Registered after completion of PET/MRI data collection (spring 2023); no statistical analysis or visualisation of outcome data before registration; blinded image processing. | Stated in Methods. |
| Primary analysis | ANCOVA (PET2 ~ PET1 + group) per outcome (VT, BPND) and region (FC, HIP); α = 0.05; no multiplicity correction. | Model 1 was prespecified in the preregistered analysis plan, while adjustment for time from dosing to PET2 (Model 2) was identified as a possible additional analysis. Model 3 included post hoc adjustments for demographic and clinical covariates. |
| Plasma free fraction | Difference in free fraction before/after psilocybin to be accounted for in VT modelling. | Plasma free fraction (fP) was not measured; therefore, potential treatment-related changes in fP could not be assessed or accounted for in VT quantification. |
| Secondary: ΔVT vs ΔMADRS (PET1→PET2) | Association between change in binding and change in MADRS. | Within-arm Pearson correlations; FDR correction, arm × ΔVT interaction, adjusted and partial-correlation models (Suppl. Tables 2–4, 8–10). |
| Secondary: baseline binding vs MADRS / ΔMADRS | Correlation between baseline binding and MADRS and ΔMADRS in all subjects. | Reported within each arm, plus arm × baseline-VT interaction (Suppl. Tables 5–6). Associations between baseline binding and baseline MADRS are reported separately. |
| Secondary: long-term outcomes (Day 42, Day 365) | ΔBinding vs long-term ΔMADRS. | As registered; observations after antidepressant reinitiation or non-protocol psychedelic use censored. |
| Exploratory questionnaires | ALIVE, AAQ-II, MEQ30, CEQ, SPIR, LEQ, MLQ, SWLS. | Reported in Suppl. Table 11. |
| Exploratory ROIs | Other ROIs may be examined exploratorily. | Extended-ROI ANCOVAs and descriptives (Suppl. Tables 12–15). |
| ROC analysis | Not registered. | Post hoc ROC analysis using the SAP-defined dual response criterion (≥50% MADRS reduction or >10-point reduction from baseline). The ≥50%-only criterion was examined in a sensitivity analysis (Supplementary Table 18). |
| Missing data | VT not estimated if critical measurements fail. | VT missing for two psilocybin participants; BPND available for 17/17 per arm. |
| Secondary biomarkers | Associations between $[^{11}C]$UCB-J binding and plasma BDNF/VEGF concentrations, and between changes in CSF serotonin-related markers, UCB-J binding and MADRS scores. | These analyses are reported separately and are not included in the present manuscript. |

Abbreviations: BDNF, brain-derived neurotrophic factor; BPND, non-displaceable binding potential; CSF, cerebrospinal fluid; FC, frontal cortex; FDR, false discovery rate; HIP, hippocampus; VT, volume of distribution; VEGF, vascular endothelial growth factor.

**Supplementary Table 17. Baseline $V_T$ and ΔMADRS at PET2 (Day 15) in the psilocybin arm: sensitivity to the participant with the highest baseline $V_T$.**

Pearson and Spearman correlations, leave-one-out range, exclusion of the highest-$V_T$ participant, and adjustment for baseline MADRS.

| ROI | Pearson r (95% CI), p; n = 15 | Spearman ρ, p; n = 15 | Leave-one-out Pearson r (range) | Pearson r (95% CI), p; excluding highest-$V_T$ participant (n = 14) | Spearman ρ, p; excluding (n = 14) | β baseline $V_T$, adjusted for baseline MADRS (p); n = 15 | β adjusted; excluding (n = 14) |
|---|---|---|---|---|---|---|---|
| Frontal cortex | −0.61 (−0.85 to −0.14), p = 0.016 | −0.39, p = 0.146 | −0.74 to −0.42 | −0.42 (−0.78 to 0.15), p = 0.138 | −0.25, p = 0.382 | −1.15 (p = 0.041) | −0.85 (p = 0.179) |
| Hippocampus | −0.65 (−0.87 to −0.20), p = 0.009 | −0.55, p = 0.035 | −0.75 to −0.49 | −0.49 (−0.81 to 0.05), p = 0.073 | −0.44, p = 0.114 | −1.53 (p = 0.034) | −1.17 (p = 0.132) |

Negative ΔMADRS values indicate symptom improvement; ΔMADRS = MADRS at Day 15 − baseline MADRS. The highest-$V_T$ participant was the same in both regions (baseline MADRS 33, ΔMADRS −30). Nominal p values; no FDR correction is applied in this sensitivity table. The Pearson correlations for all participants reproduce those in Supplementary Table 5A.

**Supplementary Table 18. Post hoc ROC analysis of baseline $V_T$ for clinical response at PET2 (Day 15): SAP-defined criterion and sensitivity analysis.**

Psilocybin arm, n = 15 with baseline $V_T$ and Day 15 MADRS.

| Response definition | Responders / non-responders | ROI | AUC (95% bootstrap CI) | AUC excluding highest-VT participant |
|---|---|---|---|---|
| SAP-defined (main): ≥50% MADRS reduction or >10-point reduction | 11 / 4 | Frontal cortex | 0.89 (0.66–1.00) | 0.88 |
| SAP-defined (main): ≥50% MADRS reduction or >10-point reduction | 11 / 4 | Hippocampus | 0.91 (0.68–1.00) | 0.90 |
| Sensitivity: ≥50% MADRS reduction only | 10 / 5 | Frontal cortex | 0.86 (0.60–1.00) | 0.84 |
| Sensitivity: ≥50% MADRS reduction only | 10 / 5 | Hippocampus | 0.88 (0.64–1.00) | 0.87 |

AUC confidence intervals were estimated by stratified percentile bootstrap (10,000 iterations; seed 42), resampling responders and non-responders separately. The main analysis uses the dual response criterion specified in the statistical analysis plan (SAP): ≥50% MADRS reduction or >10-point reduction. The ≥50%-only criterion, also used in the study protocol, is presented as a sensitivity analysis. ROC analyses were post hoc. Estimates should be interpreted cautiously given the small number of non-responders.

### 19. Supplementary Methods and Results: ROC analysis of baseline [$^{11}$C]UCB-J $V_T$

To further characterize baseline [$^{11}$C]UCB-J $V_T$ as a candidate pretreatment response marker, post hoc receiver operating characteristic (ROC) analyses were performed in the psilocybin arm. The main analysis used the response criterion specified in the statistical analysis plan (SAP): either a ≥50% reduction in MADRS total score or a >10-point reduction from baseline. For the PET2 (Day 15) endpoint, 15 participants had complete baseline $V_T$ and clinical data, of whom 11 met this criterion and 4 did not. The ROC direction was specified such that higher baseline $V_T$ predicted response. AUC 95% confidence intervals were estimated by stratified bootstrap resampling (10,000 iterations), with responders and non-responders resampled separately with replacement. A sensitivity analysis used the ≥50% reduction criterion alone (10 responders, 5 non-responders; Supplementary Table 18).

Using the SAP-defined response criterion at PET2 (Day 15), baseline $V_T$ yielded an AUC of 0.89 (bootstrap 95% CI 0.66–1.00) in frontal cortex and 0.91 (0.68–1.00) in hippocampus. Under the alternative ≥50%-only definition, corresponding AUCs were 0.86 (0.60–1.00) and 0.88 (0.64–1.00), respectively (Supplementary Table 18). Given the small sample, particularly the four non-responders under the SAP criterion, these exploratory estimates are uncertain and require prospective external validation.

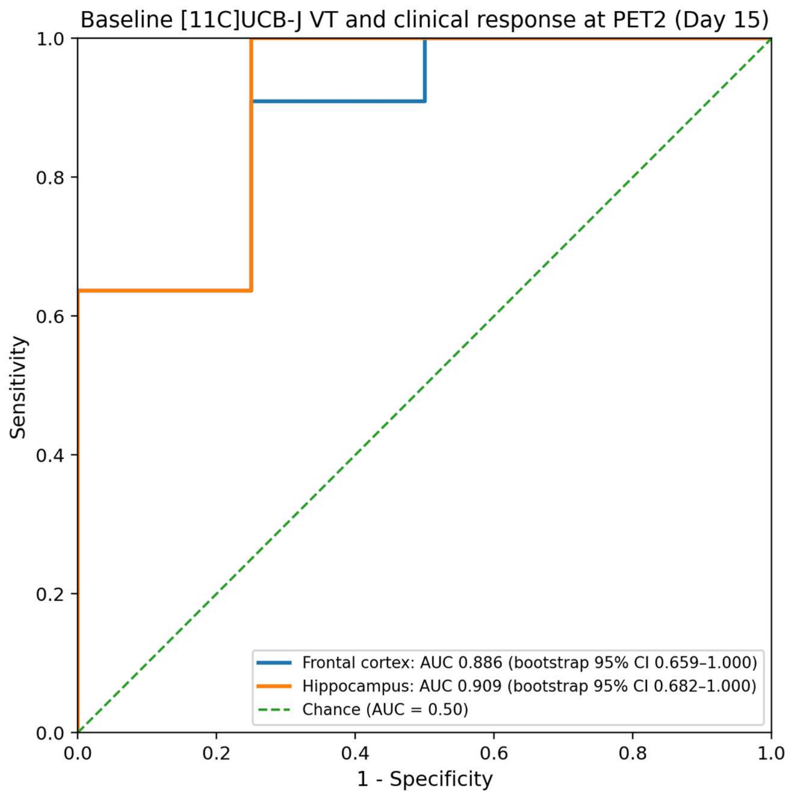


**Supplementary Figure 1.** Baseline [$^{11}$C]UCB-J $V_T$ as a classifier of clinical response at PET2 (Day 15) in the psilocybin arm. Response was defined using the statistical analysis plan (SAP) criterion: ≥50% reduction in MADRS total score or >10-point reduction from baseline (11 responders, 4 non-responders). ROC curves are shown for frontal cortex and hippocampus; labels report AUC estimates with 95% confidence intervals derived from 10,000 stratified bootstrap iterations. The diagonal dashed line denotes chance-level discrimination (AUC = 0.50).
